# Patients’ and carers’ experiences of, and engagement with remote home monitoring services for COVID-19 patients: a rapid mixed-methods study

**DOI:** 10.1101/2021.12.17.21267968

**Authors:** Holly Walton, Cecilia Vindrola-Padros, Nadia Crellin, Manbinder S Sidhu, Lauren Herlitz, Ian Litchfield, Jo Ellins, Pei Li Ng, Efthalia Massou, Sonila M Tomini, Naomi J Fulop

**Affiliations:** Department of Applied Health Research, University College London, Gower Street, London WC1E 6BT, UK; Department of Targeted Intervention, University College London, Charles Bell House, 43-45 Foley Street, London, W1W 7TY; Nuffield Trust, 59 New Cavendish St, London W1G 7LP; Health Services Management Centre, School of Social Policy, College of Social Sciences, University of Birmingham, 40 Edgbaston Park Rd, Birmingham, B15 2RT, UK; Institute of Applied Health Research, College of Medical and Dental Sciences, University of Birmingham, 40 Edgbaston Park Rd, Birmingham, B15 2RT, UK; Health Services Management Centre, University of Birmingham, Park House, 40 Edgbaston Park Road, Birmingham, B15 2RT; Department of Public Health and Primary Care, University of Cambridge, UK

**Keywords:** Remote home monitoring, COVID-19, patient experience, patient engagement, care

## Abstract

**Introduction:** Remote home monitoring models were implemented during the COVID-19 pandemic to shorten hospital length of stay, reduce unnecessary hospital admission, readmission and infection, and appropriately escalate care. Within these models, patients are asked to take and record readings and escalate care if advised. There is limited evidence on how patients and carers experience these services. This study aimed to evaluate patient experiences of, and engagement with, remote home monitoring models for COVID-19.

**Methods:** A rapid mixed-methods study in England. We conducted a cross-sectional survey and interviews with patients and carers. Interview findings were summarised using rapid assessment procedures sheets and grouping data into themes (using thematic analysis). Survey data were analysed using descriptive statistics.

**Results:** We received 1069 surveys (18% response rate) and conducted interviews with patients (n=59) and carers (n=3). ‘Care’ relied on support from staff members, and family/friends. Patients and carers reported positive experiences and felt that the service and human contact reassured them and was easy to engage with. Yet, some patients and carers identified problems with engagement. Engagement was influenced by: patient factors such as health and knowledge, support from family/friends and staff, availability and ease-of-use of informational and material resources (e.g. equipment), and service factors.

**Conclusion:** Remote home monitoring models place responsibility on patients to self-manage symptoms in partnership with staff; yet many patients required support and preferred human contact (especially for identifying problems). Caring burden and experiences of those living alone, and barriers to engagement should be considered when designing and implementing remote home monitoring services.

**Patient or public contribution:** For this evaluation, members of the study team met with service user and public members of the BRACE PPI group and Health and Care Panel and patient representatives from RSET in a series of workshops. These workshops informed study design, data collection tools, data interpretation and to discuss study dissemination for Phase 2. For example, patient facing documents, such as the consent form, topic guides, patient survey and patient information sheet were reviewed by this group. Additionally, PPI members helped to pilot patient surveys and interview guides with the research team. We also asked some members of the public to pilot the patient survey. Members of the PPI group were given the opportunity to comment on the manuscript. One PPI member commented on the manuscript and the manuscript was amended accordingly.

## Introduction

In recent years, there has been a shift in healthcare delivery [1]; with services having adopted technology in different ways, including virtual consultations [2-5], or remote monitoring models of healthcare [1,5]. Within remote home monitoring models, patients and carers are asked to record health readings in one place (e.g. at home), and these readings are reviewed and responded to by professionals elsewhere [6-7]. These changes in healthcare delivery potentially alter the landscape of ‘care’, as they accompany or even move away from traditional face-to-face care models [8], and instead place further emphasis on formal or informal carers providing care at a distance and reviewing readings remotely [9].

This shift in healthcare delivery is also consistent with recent moves towards self-management and patient activation within healthcare, whereby accountability for care has changed [10-12]. Patients are becoming more involved in self-management, for example, learning how to detect and manage their symptoms, and treatments, and escalation of care associated with their condition [7,13-17], and healthcare tasks (e.g. managing medication, organising care appointments, taking measurements) [18]. Whilst some patients may welcome this [19], there have been concerns that self-management puts burden onto patients and families, rather than facilitating shared care [10, 19]. Additionally, the effectiveness of these concepts is not fully understood yet [12,19-23].

The COVID-19 pandemic further enhanced and accelerated the need for healthcare services to use technology in care delivery [5] and escalated the need for patient self-management. Remote home monitoring models have previously been used to provide care for chronic conditions [24-26]. During the pandemic, remote home monitoring models were used for acute conditions such as COVID-19, with the aim to shorten length of stay in hospital, reduce unnecessary hospital admissions or readmission and infection transmission, and escalate care as needed [27-28].

Many different types of COVID-19 remote home monitoring models were implemented throughout England. Some models referred patients from community services (e.g. GPs, hot hubs, and emergency departments), known as COVID Oximetry @home [27]. Others referred patients onto the service as early discharges from hospital, known as COVID virtual wards [28]. See Box 1 for a brief description of services [27-29].

### Box 1.

Description of COVID-19 remote home monitoring services [27-29]

- Patients are given a pulse oximeter, together with information and resources outlining how to use the equipment, escalation warning signs and what to do if these warning signs appear.
- Patients measure their oxygen saturation levels using the oximeter and other readings (pulse/heart rate/temperature) regularly and record and submit these readings. Readings are shared by telephone or using a tech-enabled method (e.g. an app on the patient’s phone or computer).
- Patients are then escalated for further care if necessary.
- Discharge from the service is typically around 14 days

Whilst remote home monitoring models may reduce the need for staff to assess patients in person, they place more responsibility, commitment and workload onto patients and carers [10]. For example, in COVID-19 remote home monitoring services, patients and carers are expected to measure and record oxygen saturations, and escalate care if readings drop below certain thresholds [29-30]. This increased responsibility may be appropriate and beneficial for some patients but may not be suitable for everyone [31]. Some people may be unable to meet expectations placed on them by healthcare services and experience negative impacts from treatment burden [10]. Negative impacts may include health consequences faced by patients due to not adhering to treatment and on patients’ professional, social, emotional and financial situation [18]. Different individuals may tolerate different levels of treatment burden, and it has been suggested that this needs to be assessed regularly as tolerance changes over time [10,32]. Many factors worsen treatment burden, including: situational factors (e.g. travel), personal factors (e.g. beliefs and relationships), and structural factors (e.g. treatment factors and access to resources) [18]. Therefore, formal and informal support networks are needed to support patients [7,33].

Treatment burden may negatively impact on patient experience and levels of engagement. This is problematic given that patient engagement with remote home monitoring is crucial. Patient engagement has been defined as patients understanding the information they are given (‘receipt’) and being able to perform the required activities (‘enactment’) [34-35].

Whilst previous research indicates factors which may influence patient engagement with treatment models more generally [7,10,18,33], there is a lack of research on patient experience and engagement with remote home monitoring services for an acute condition such as COVID-19 [29-30]. If patients do not engage with these services, they may be at risk of negative outcomes that the service aimed to prevent e.g. silent hypoxia (very low oxygen saturations, often without breathlessness) [36] and/or delayed admission to hospital [37-38]. Additionally, if engagement is limited then it is not possible to evaluate whether or not the service influences key outcome measures such as any changes in mortality or hospital use. This research addresses this gap by evaluating patient experience of, and engagement with COVID-19 remote home monitoring services.

This study aimed to explore what formal and informal support patients received as part of COVID-19 remote home monitoring services (COVID Oximetry@home and virtual wards models), and patient experience of and engagement with these services. This manuscript addressed the following questions:

1. What types of formal and informal support did patients receive as part of COVID-19 remote home monitoring services? What was the burden of treatment on patients and carers in informal support roles?
2. What are patients’ and carers’ experiences of engaging with COVID-19 remote home monitoring services?
3. What are the factors influencing burden of treatment and ability to engage with COVID-19 remote home monitoring services?

## Methods

### Design

This manuscript draws on cross-sectional survey data from patients and carers and qualitative data from semi-structured interviews with patients and carers. This was a rapid study (data collection period: March-June 2021). The methods are reported in detail in Appendix 1.

This study was part of a larger rapid mixed-methods evaluation of remote home monitoring for COVID-19 patients [39].

### Sample

We recruited patients and carers from 25 sites (COVID-19 remote home monitoring services). Sites were sampled using a range of criteria (e.g. the mechanism for patient monitoring and their geographic location). We recruited sites from across eight English regions, and these covered populations of <250,000 to over 1 million (see Appendix 2 for details). Seventeen of the 25 sites participated in both surveys and in-depth interviews, the remaining sites were survey only sites.

Patients who had received COVID-19 remote home monitoring services were recruited to the survey (aimed to recruit all onboarded patients between January 2021 and June 2021) and the interviews (4-6 patients/carers from each of the 17 case study sites). If patients were unable to take part but wanted to participate, we invited their carer to complete the survey/interview on their behalf.

### Measures

We developed the survey and semi-structured topic guides specifically for this study. Questions were informed by relevant service documentation [27-28], theoretical frameworks relating to social, political and technical contexts [7,8,33,40] and behaviour [41], and previous literature on engagement [34-35]. The patient topic guide and survey covered questions relating to patients’ experiences of, and engagement with the service, including things that helped and got in the way, recommendations to improve the service, and demographic characteristics (e.g. gender, age, education, ethnicity, work, living situation, health, and sexuality) [42-47] (see Appendix 3-4). Information sheets and the survey were available in six other languages (Polish, Bengali, Urdu, Punjabi, French and Portuguese).

The survey and interview guide were piloted with the members of the study public patient involvement (PPI) group and the general public, through the following activities: a) workshop with PPI group, b) pilot interview with one PPI member, c) survey reviewed by PPI member and members of public. Suggested amendments relating to accessibility, and wording of questions were incorporated prior to use.

### Data collection

Study coordinators working within each service distributed electronic or paper surveys to patients and carers.

Potential interview participants were approached by study coordinators from each site. If they were interested in taking part, they were contacted by a researcher who sent them an information sheet and consent form. Participants were asked to return the consent form before the interview. Interviews were conducted by six researchers. Interviews were conducted over Microsoft Teams, Zoom or telephone.

### Analysis

Survey data were analysed using SPSS statistical software (version 25). Descriptive statistics were used to explore patient experience and engagement. For data relating to patient experience and engagement, all cases were analysed (whether carer, patient or unknown). Where data were missing for specific questions, cases were excluded from the analysis and the denominator reported. Open text survey data were extracted into an Excel spreadsheet and coded inductively. We extracted data from three questions: if patients had anything else to say about the service (n=434 open text responses), how carers have supported their friend/family while they had COVID-19 (n=61 open text responses) and recommendations to improve the service (n=200 open text responses).

Interview data were analysed using Rapid Assessment Procedure (RAP) sheets. RAP sheets are tools which can be used to rapidly capture key findings from different data sources [48]. Research leads from each site added notes and summaries of findings to the RAP sheet following each interview, for each site. The data inputted into RAP sheets were inductively coded using thematic analysis by one researcher (HW). Themes and sub-themes were developed, discussed and agreed by the research team. We then developed a framework based on these themes and sub-themes and one researcher (HW) used this framework to extract quotes from all original transcripts. The coding framework included: participants’ views of the service, experiences of being referred, information received about the service and experiences performing remote home monitoring behaviours, and barriers and facilitators to performing remote home monitoring behaviours.

Survey and interview findings were triangulated.

## Results

### 1. Participant characteristics

We received 1069 surveys (18% response rate) from patients (n=936, 87.6%) and carers (n=48, 4.5%) across 25 sites (see Appendix 5). In some surveys it was unclear whether it was completed by the patient or the carer (n=85, 8%). We conducted 62 interviews: with patients (n=59) and carers (n=3) across 17 sites (see Appendix 6 for demographics). Our sample of CO@h patients were generally representative of CO@h patients that were onboarded nationally, with under-representation and over-representation of some groups [49].

Most patients were referred to the service via community methods (see Table 1). Patients and carers reported using a range of methods to record and report their readings to the service, including analogue (paper and phone) and tech-enabled methods (see Table 1).

**Table 1.**
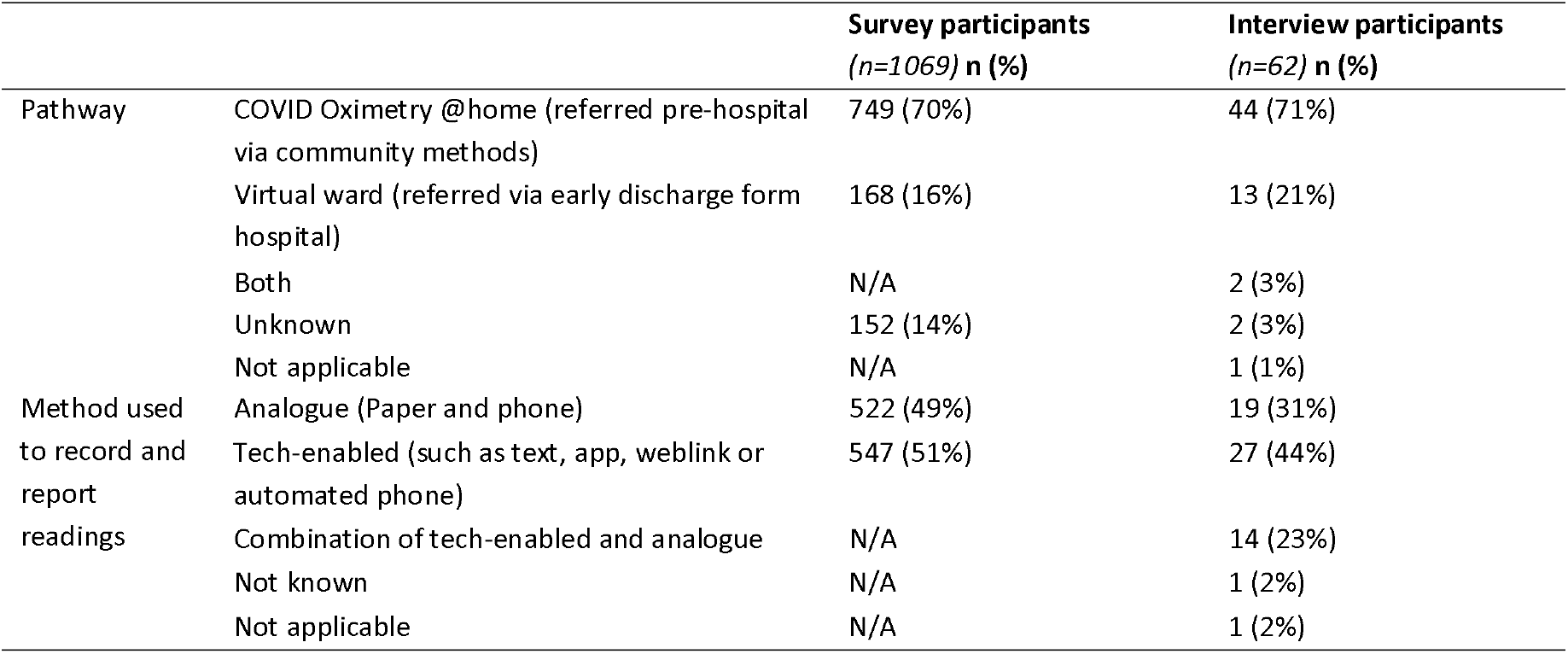
Summary of patients’ remote home monitoring pathway and method of recording and reporting

|  |  | Survey participants<br>(*n*=1069) *n* (%) | Interview participants<br>(*n*=62) *n* (%) |
| --- | --- | --- | --- |
| Pathway | COVID Oximetry @home (referred pre-hospital via community methods) | 749 (70%) | 44 (71%) |
|  | Virtual ward (referred via early discharge form hospital) | 168 (16%) | 13 (21%) |
|  | Both | N/A | 2 (3%) |
|  | Unknown | 152 (14%) | 2 (3%) |
|  | Not applicable | N/A | 1 (1%) |
| Method used to record and report readings | Analogue (Paper and phone) | 522 (49%) | 19 (31%) |
|  | Tech-enabled (such as text, app, weblink or automated phone) | 547 (51%) | 27 (44%) |
|  | Combination of tech-enabled and analogue | N/A | 14 (23%) |
|  | Not known | N/A | 1 (2%) |
|  | Not applicable | N/A | 1 (2%) |

### 2. What types of formal and informal support did patients receive as part of COVID-19 remote home monitoring services?

#### Formal support from staff

The ‘care’ on offer differed across sites and patients; with variation in the type and frequency of monitoring offered by services. Our wider evaluation indicated that all the services offered patients support for monitoring and escalation (with varying intensity) [49].

Responses from the patient survey indicated that the frequency with which patient’s had contact with a member of staff ranged from several times a day to not at all. Most patients and carers had contact either once a day, or several times a week. A few patients and carers reported not speaking to staff at all (see Table 2). This is supported by interview findings which indicated that the frequency of taking and communicating readings to the service ranged from once a day to more than three times a day. Findings indicate that patients are supported by staff throughout different stages of the service, including providing information, monitoring (e.g. phone calls if patients and carers forget to submit readings and in some cases face-to-face visits to take readings), escalating care (e.g. providing advice on whether to seek help, calling ambulances for patients), signposting and comfort and reassurance.

**Table 2.** Summary of frequency of contact with staff members for survey participants

| **Frequency of contact with staff member** | **Percentage of survey participants n (%)** | **Percentage range across sites** |
| --- | --- | --- |
| Several times a day (*n=1060*) | 169 (16%) | 0%-45% |
| Once a day (*n=1060*) | 276 (26%) | 7%-91% |
| Several times a week (*n=1060*) | 270 (25%) | 0%-62% |
| Once a week (*n=1060*) | 139 (13%) | 0%-31% |
| Less than once a week (*n=1060*) | 142 (13%) | 0%-27% |
| Not at all (*n=1060*) | 64 (6%) | 0%-27% |

#### Burden of treatment on patients and carers in informal support roles

Survey findings indicated that almost all patients used an oximeter to record readings when receiving the service. Many patients reported completing a diary and providing readings over the phone or using technology-enabled methods. Escalation related behaviours were reported less frequently by patients, with only a third of patients reporting seeking further help due to readings being lower than recommended thresholds, and only a fifth of patients checking their readings for issues (see Table 3).

**Table 3.** Remote home monitoring activities reported by survey participants

| **Remote home monitoring activities that patients reported doing** | **Percentage of survey participants n (%)** | **Percentage range across sites** |
| --- | --- | --- |
| Using the oximeter (*n=1069*) | 1014 (95%) | 81%-100% |
| Completing a diary (*n=1069*) | 555 (52%) | 11%-89% |
| Providing readings over the phone (*n=1069*) | 498 (47%) | 19%-93% |
| Providing readings via text (*n=1069*) | 309 (29%) | 0%-74% |
| Recording readings in a digital app (*n=1069*) | 264 (25%) | 0%-89% |
| Providing readings via email (*n=1069*) | 15 (1%) | 0%-5% |
| Seeking further help due to readings being lower than the recommended threshold (*n=1069*) | 344 (32%) | 18%-71% |
| Checking over readings for issues (*n=1069*) | 215 (20%) | 7%-41% |

Many patients were supported by family and friends to engage with the service. A quarter of survey respondents needed help to use equipment (25%, range 11%-50% across sites), and more than half of the interview participants were supported by family members.

Most patients and carers reported having informal support to help them use the oximeter and support with taking and recording readings. Only a small proportion of participants reported that they did not need support using the oximeter (10%) or taking and recording readings (19%) (see Table 4). Qualitative findings highlighted that family and friends provided support with the following activities: support submitting readings or communicating readings over the phone, support with monitoring, support collecting oximeter, support writing down readings and contacting and taking calls from the service, translation support or using the app. Additionally, other patients reported that their family members and friends provided comfort and reassurance, support as and when needed, support and advice at a distance, and domestic care. Open text survey responses indicated that many carers provided full time care for their family member/friend whilst on the service. Some carers were family members or friends who moved in to provide support. Others made regular telephone calls to check in on their family member.

**Table 4.** Support for remote home monitoring activities as reported by survey participants

| Support for remote home monitoring activities |  | Percentage of survey participants<br>n (%) | Percentage range across sites |
| --- | --- | --- | --- |
| Having someone to help use the oximeter when needed (*n*=1058) | Yes | 782 (74%) | 50%-100% |
|  | No | 169 (16%) | 0%-44% |
|  | Not applicable | 107 (10%) | 0%-22% |
| Support taking and recording readings if needed (*n*=1057) | Yes | 666 (63%) | 43.8%-90% |
|  | No | 190 (18%) | 7%-33% |
|  | Not applicable | 201 (19%) | 0%-37.5% |

However, not all patients had support with using the oximeter (16%) or taking/recording readings (18%). Some patients did not have support due to their family members having COVID-19.

### 3. What are patient and carers’ experiences of engaging with COVID-19 remote home monitoring services?

Patients mostly had positive views of the service. 93% (n=970/1045) of survey respondents rated the service as excellent or good, 90% (n=923/1028) of respondents found the service helpful and 91% (n=944/1037) would recommend the service to their family and friends.

Findings indicated that most patients and carers found the service reassuring and supportive (91% (n=946/1040) of survey respondents). Qualitative findings indicated that patients and carers valued the human contact with staff and found it reassuring due to having someone watching over them; particularly for those who were living alone, had no support nearby or had existing conditions.

> *“Because it’s obviously keeping an eye on you, isn’t it really? And I was getting the phone calls every day. How are you feeling?, […]. But someone who was on their own, who had no-who was living on their own, you know, it’s a bit of a lifesaver isn’t it?”* (Site A, interviewee 4)

A minority of patients and carers felt that there were gaps in the service, and it was not holistic. Some felt that the service was narrowly focused on managing known symptoms of COVID-19 which did not always suit those with other symptoms, health conditions or who required wider support. A few patients reported feeling that the service was isolating and unsupportive (e.g. they only received a call about the oximeter drop off/return, but not for monitoring).

Most patients and carers felt the care provided was appropriate and preferred to be at home instead of being in hospital (given the pandemic context). Reasons for preferring home over hospital included freeing up space for others in need, being familiar with your environment, fears of going to hospital during a pandemic, communication barriers in hospital, being able to work and perceptions that home monitoring was a suitable care package for those with more minor symptoms of COVID-19. However, some patients and carers spoke about preferring to be in the hospital rather than at home, to feel more secure, feeling scared and wanting to be seen face-to-face.

> *“And I didn’t feel too embarrassed that I was using up valuable resources because I thought, Well I’m sitting here at home, there’s no reason for me to go in Hospital, bother anyone and waste people’s time*.*”* (site N, interviewee 3)

A minority of patients and carers spoke about how the service was the only available care and that they would have liked to have received care from other healthcare professionals such as their GP in parallel. Many patients and carers were not aware of the service prior to referral.

Some patients and carers also spoke about how the service helped them to monitor their own improvement and that it potentially improved their outcomes.

Patients and carers reported very positive views of the workforce and that they were helpful and put patients at ease and were professional and potentially even lifesaving. Continuity of staff was thought to be important.

A few patients had negative experiences with individual staff members e.g. that they were dismissive, did not recognise that they needed help, were not interested or lacked clinical expertise to support patients or answer their queries.

### 4. What are the factors influencing burden of treatment and ability to engage with COVID-19 remote home monitoring services?

Findings indicated that patients and carers generally found it easy or very easy to engage with the service and the resulting activities, including understanding information, monitoring using the oximeter, recording readings and providing readings and escalating care (see Table 5). Most survey respondents indicated that they did not experience problems with the service (72%; n=771/1069) and did not report barriers to engagement with the service (80%, n=858/1069).

**Table 5.** Experience engaging with service activities as reported by survey participants

| **Experience engaging with the service activities** | **Percentage of survey participants who reported easy/very easy n (%)** |
| --- | --- |
| Understanding the information they were given (n=1040) | 970 (93%) |
| Monitoring using the oximeter (n=1049) | 1022 (97%) |
| Recording readings (n=949) | 913 (96%) |
| Providing readings to the remote home monitoring team (n=1010) | 979 (97%) |
| Seeking further help (if applicable) (n=857) | 738 (86%) |

Engagement with service activities was not without challenges, with some patients and carers reporting issues with the information provided or needed further information. Some patients found monitoring difficult due to other health conditions, or that monitoring made them feel worried. Some patients and carers wanted more support or found recording burdensome. Finally, some issues related to escalating care were identified in the interviews. The uncertainty of COVID-19, perceptions of hospital as a frightening places and uncertainty around interpretation of readings and thresholds, meant that some patients and carers were hesitant to self-escalate their care, waited for a member of staff to advise them to escalate their care or reported not wanting to go to hospital or seek further support even when advised to by staff members.

> *“really I should have probably rung when the readings were that bad but I didn’t. […]And when I did send them through they said, ‘No, get to the doctors now’*.*” (Site C interviewee 6)*

The most frequent challenges reported within the survey were returning the oximeter, contacting healthcare professionals when needed and seeking further help (see Table 6). Whilst many survey respondents discussed problems with the team (35%, n=87/249) or had their problems resolved (33%, n=76/232), over half of these participants said problems had not been resolved (54%, n=126/232).

**Table 6.**
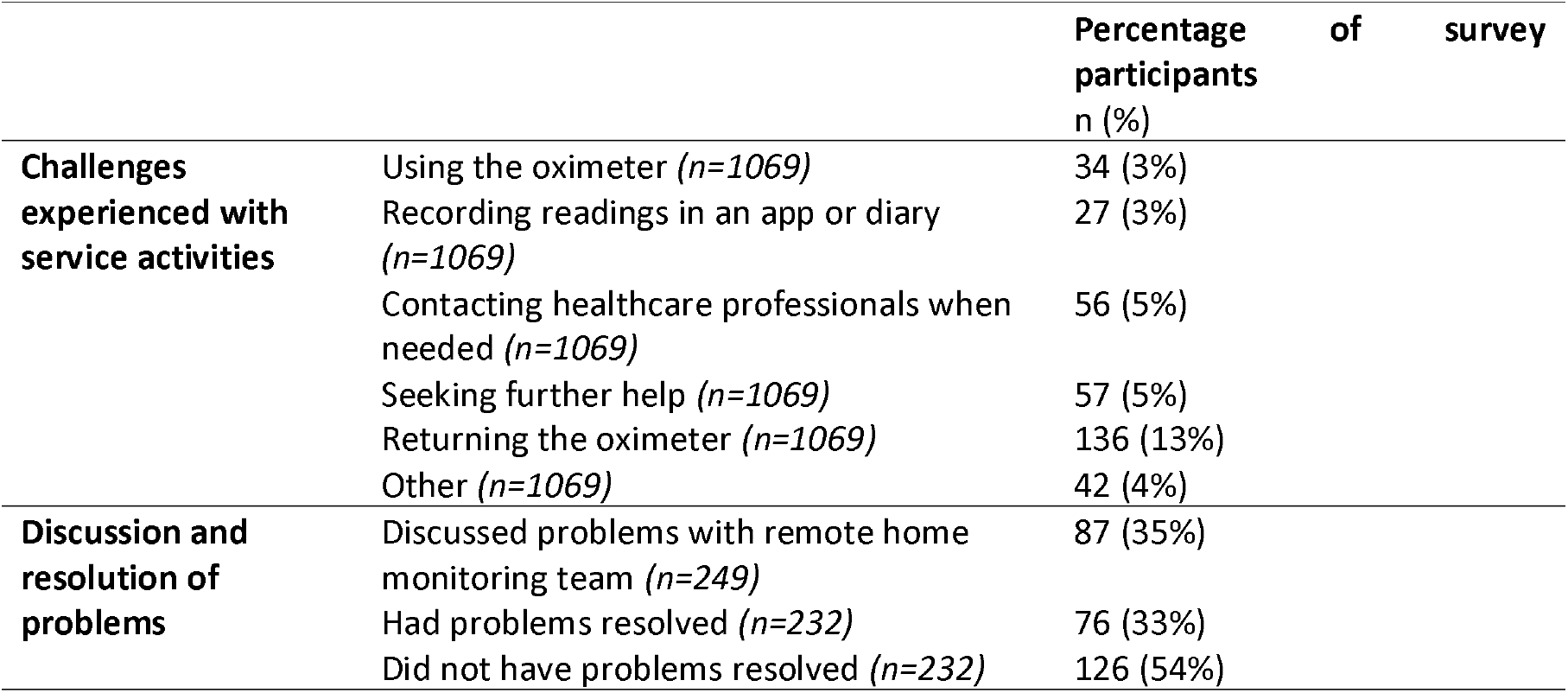
Challenges experienced and the discussion and resolution of problems as reported by survey participants

Findings from the surveys and interviews indicated three overarching themes that influenced burden of treatment and patient’s ability to engage, with COVID-19 remote home monitoring services: i) patient factors, ii) wider support and resources, and iii) factors relating to the service (see Appendix 7 for details of example findings for each theme and sub-theme and example quotes).

#### Patient factors

Knowledge, memory, physical health, attitudes towards the service and having time to complete the required tasks influenced engagement (see Appendix 7). Interview findings indicated that patients in poorer health (e.g. due to COVID-19, other health conditions) found it harder to engage with the service. For example, many participants spoke about feeling too unwell due to COVID-19 (often during the first few days of the service) and therefore they were unable to engage with monitoring behaviours such as taking and recording readings. Some patients and carers spoke about having other health conditions which made it difficult to engage with monitoring behaviours (e.g. hearing and eyesight difficulties). Patients and carers who felt they had sufficient knowledge about what they needed to do found it easier to engage (e.g. 55% (n=583/1069) of survey respondents felt that knowing what to do helped them to engage with the service). On the other hand, a lack of knowledge on how to complete the activities (e.g. a lack of knowledge on how to escalate care or what the thresholds for escalating care are) limited engagement.

#### Wider support and resources

Support from staff/service, support from family members/friends, accessibility and availability of materials, equipment and technology influenced engagement (see Appendix 7). For example, support from staff members (e.g. 46% (n=488/1069) of survey respondents) and family/friends (e.g. 25% (n=266/1069) of survey respondents) was crucial in helping many patients to use the service.

#### Service factors

Monitoring characteristics, service characteristics, scope of service and availability of treatment influenced engagement (see Appendix 7). For example, some participants felt that the inconsistent timing of calls was a barrier and some felt that calls were too frequent, whereas others felt they were not frequent enough. Additionally, some patients and carers felt that the scope of the service was a barrier to engagement; in that it did not cover wide symptoms of COVID-19 and was not holistic.

## Discussion

### Key findings

Findings indicated that patients can engage with remote home monitoring services, even when experiencing acute illnesses (e.g. COVID-19). However, many patients required formal input from staff and informal support from family and friends to complete the necessary tasks (e.g. monitoring oxygen saturations). Patients and carers had positive experiences receiving remote home monitoring. The human contact from staff provided patients and carers with reassurance and patients reported the service was mostly easy to engage with. However, patients’ ability to engage with the service was conditional on a range of factors, including having support from family/friends and staff, being in good health and receiving clear instructions on what they needed to do and how to do it, and the level of commitment from patients whilst on the service.

### How findings relate to previous research

Earlier studies have explored types of remote home monitoring, and implementation of and cost of remote home monitoring models for COVID-19 [29-30]. Yet there was little research on patient experiences of remote home monitoring services when delivered during a pandemic; in the context of pressured health services and concerned patients. These findings extend earlier findings by highlighting patients’ and carers’ positive views of the service, challenges and concerns relating to engagement with remote home monitoring for acute conditions, and tangible recommendations on how to improve remote home monitoring services (see Appendix 8); many of which support wider themes reported in patient experience literature (e.g. the importance of information provision) [50-51].

Previous research outlines concerns relating to remote care and telemedicine and the loss of interpersonal dimensions involved in caring relationships [9]. Findings extend the evidence base by showing that care does not need to take place face-to-face for patients to feel reassured and supported. Patients largely felt care provided at a distance was appropriate and that they were being monitored. This is consistent with previous research indicating that technology may support closer contact with professionals [9]. However, findings may have been affected by the pandemic context in that data was collected during the height of wave 2 of the pandemic, therefore patients may have been more likely to accept remotely delivered services to help minimise risk to themselves, family members and staff. Patients and carers may feel differently about remote home monitoring and care delivered at a distance in non-pandemic contexts.

New models of healthcare such as COVID-19 remote home monitoring services sought to change the traditional model of in person care. Instead, within these models, staff engage with patients to share the care burden whilst equipping patients and carers to self-monitor and manage care in the absence of staff members. Our findings demonstrate that concepts of treatment burden and difficulties engaging with healthcare demands [10, 18, 32] also apply to remote monitoring models for COVID-19. Some patients reported problems engaging with remote home monitoring services for a range of reasons, including feeling too poorly, not having enough knowledge on what to do, and lack of support from staff and/or family/friends. Others reported the necessity of support from their family/friends when engaging with the service. This extends knowledge by showing that social networks may undertake self-monitoring tasks on behalf of (potentially very poorly) patients in addition to helping patients cope with burden of treatment [10] and self-management of conditions [7, 52]. Those who feel more poorly (either due to COVID-19 or existing conditions) may require more formal or informal support to manage care. Yet, this increases the caring burden for family/friends. This finding also raises concerns regarding appropriateness of care for those who do not have informal support networks in place.

This manuscript builds on earlier research by providing a nuanced interpretation of the factors that influenced engagement with remote home monitoring services for acute conditions such as COVID-19. Our finding that many non-health related factors influence engagement is consistent with other studies in a range of other conditions and interventions (7, 18, 53-56). Our findings add to prior knowledge by demonstrating that many factors were exacerbated due to the acute nature of COVID-19 and policy factors surrounding COVID. For example, physical health factors limiting engagement may be worsened by the acute nature of COVID-19 and the severity of symptoms that some patients faced, thus affecting a patient’s ability to engage. Furthermore, policy regulations and lockdown restrictions imposed within the UK may have meant that physical social support available to patients may have been limited by members of their support networks living elsewhere. As COVID-19 is easily transmissible, social distancing recommendations were in place, therefore, many patients were distancing from family members living in the same space. Findings demonstrate that despite difficulties imposed by COVID-19, social networks were crucial for many patients in facilitating engagement with the service and ensuring that care needs were met. This highlights the need for alternative support where necessary (particularly for those living alone or those who are socially isolated). The reliance on informal support networks has implications for burden of treatment and may not be appropriate for all individuals. These findings support previous theoretical frameworks indicating that social, political and technical contexts influence engagement [7,8,33,40].

Previous research has explored concepts of self-management [7,33,53], engagement [54] and treatment burden [18] in chronic conditions, but little research had been conducted on remote home monitoring and self-management in acute conditions such as COVID-19; in which care needs to be urgently escalated in an efficient and time sensitive manner. Findings extend earlier work by demonstrating challenges of remote home monitoring models for acute conditions. For example, due to uncertainty of COVID-19, perceptions of hospital being a frightening place and uncertainty around readings and thresholds, some patients were hesitant to self-escalate care and in many cases, patients waited until advised by staff to escalate care. This finding indicates that in situations where there is a need for timely escalation, but concerns around infection transmission from going to hospital, it may be suitable to have formal support from staff members. Together, staff and patients can collaboratively decide when to seek further help, rather than placing responsibility onto patients. This finding contrasts with recommendations within the national standard operating procedures for COVID-19 remote home monitoring services [27-28], which indicate that pathways should encourage patients to self-escalate care.

Some patients felt that they were able to engage with the service due to having manageable symptoms and feeling comfortable with tasks. This indicates different levels of remote monitoring support are needed for different individuals. This supports previous research indicating that the success of telehealth services including remote home monitoring may rely on the fit between individuals’ needs and services [9,33].

### Strengths and limitations

Integration of mixed-methods data helped to provide in-depth perspectives on experiences of, and engagement with, COVID-19 remote home monitoring services. A large team of researchers (from a range of disciplines, with extensive expertise in qualitative and quantitative methods) were involved, thus strengthening interpretation of findings. Findings were shared with clinical and academic stakeholders. Our study sampled a large range of sites with a range of characteristics, thus enhancing generalisability of findings.

Compared with patient onboarding data, our patient sample was under-representative of some groups (e.g. older patients, Black, Asian and minority ethnic (BAME) communities and most deprived) and over-representative of other groups [49]. The response rate for the survey was fairly low (17.5%). Additionally, we were unable to recruit interview or survey participants who had declined the service, dropped out from the service, and those who were unable or did not want to take part in surveys and interviews. Therefore, findings may not be representative of all patient groups and experiences.

While we did include carers within our sample, the focus of our research as on patient experiences of remote home monitoring services. Therefore, it is possible that we have not captured carers’ experiences in detail. However, some carers shared their own experiences during the interviews and in responding to the survey.

### Implications

Burden of treatment may not only affect those with multimorbidity or chronic conditions, but can also affect those with acute conditions. Findings indicate that remote monitoring may increase treatment burden for some patients and families.

COVID-19 remote home monitoring services aimed to target patient groups at higher risk from COVID-19, yet many of these groups appear more likely to report difficulties in engagement with these services, e.g., older patients, and patients with health problems. Remote monitoring may not be appropriate for everyone (e.g., those without support). Services need to gauge a person’s support network and any concerns surrounding remote home monitoring when assessing eligibility for these services. Services must then tailor the healthcare offer to enable patients to engage (e.g. providing further support for those from at-risk groups or who do not have informal support, or linking patients with care networks if needed). All patients should be contact details to contact the service should problems arise. Face-to-face support (e.g. for monitoring) from staff and families has implications for infection transmission.

Our findings may have implications for remote home monitoring services more generally. Service developers should consider the type of condition when designing pathways. For example, services for acute conditions may require support from staff to ensure that patients are escalated for further care as necessary. Services must plan logistics for delivery and collection of equipment, ensure sufficient information provision and that patients know what they need to do and that they feel able to engage with the service. Some patients felt that the service offered was too narrow and does not consider wider social, emotional or condition related needs. Service adaptations may be necessary for those receiving remote home monitoring for acute conditions in addition to care for other chronic conditions.

### Future research

Further research is needed to explore the experiences of those who decide not to use remote home monitoring services, or disengage from these services. Further research should explore the burden of treatment for chronic conditions compared with acute conditions. Additionally, it would be helpful to further explore which groups are able to tolerate burden associated with remote home monitoring pathways, and the impact of treatment burden from informal caring responsibilities on families.

## Conclusions

COVID-19 remote home monitoring services place a large responsibility on patients and carers in relation to monitoring and escalating care. Whilst patients and carers found the service reassuring and a positive experience, many factors influenced their ability to engage with the service. This indicates that the service may be conditional on a range of factors relating to the patient (e.g. knowledge and memory), their support and resources (e.g. support from family, friends and staff) and service factors (e.g. scope of the service and frequency of monitoring).

## Data Availability

The datasets generated and/or analysed during the current study are not publicly available due to participant confidentiality.

## Acknowledgements

We are indebted to all of the services who participated in this study and to all of the patients and carers who participated in our surveys and interviews. Thank you to the following: Dr Jennifer Bousfield for supporting with study design and data collection, Simon Barnes for supporting with data entry; Steve Morris, Chris Sherlaw-Johnson, Theo Georghiou, and Jon Sussex for advice given throughout the project; our NIHR BRACE and NIHR RSET public patient involvement members for feedback throughout the study and to Raj Mehta for commenting on a draft of the manuscript; the NIHR 70@70 Senior Nurse research Leaders for providing feedback on the development of our study; Russell Mannion for peer-reviewing our study protocol; and the NIHR Clinical Research Networks for supporting study set up and data collection.

We thank the NHS Digital CO@h Evaluation Workstream Group chaired by Professor Jonathan Benger for facilitating and supporting the evaluation, and to the other two evaluation teams for their collaboration throughout this evaluation: i) Institute of Global Health Innovation, NIHR Patient Safety Translational Research centre, Imperial College London and ii) the Improvement Analytics Unit (Partnership between the Health Foundation and NHS England and NHS Improvement).

Many thanks to our Clinical Advisory Group for providing insights and feedback throughout the project (Dr Karen Kirkham (whose previous role was the Integrated Care System Clinical Lead, NHSE/I Senior Medical Advisor Primary Care Transformation, Senior Medical Advisor to the Primary Care Provider Transformation team), Dr Matt Inada-Kim (Clinical Lead Deterioration & National Specialist Advisor Sepsis, National Clinical Lead - Deterioration & Specialist Advisor Deterioration, NHS England & Improvement) and Dr Allison Streetly (Senior Public Health Advisor, Deputy National Lead, Healthcare Public Health, Medical Directorate NHS England).

## References

[1] Braithwaite J, Vincent C, Garcia-Elorrio E, Imanaka Y, Nicklin W, Sodzi-Tettey S, Bates DW. Transformational improvement in quality care and health systems: the next decade. BMC medicine. 2020 Dec;18(1):1–7. https://doi.org/10.1186/s12916-020-01739-y

[2] Greenhalgh T, Shaw S, Wherton J, Vijayaraghavan S, Morris J, Bhattacharya S, Hanson P, Campbell-Richards D, Ramoutar S, Collard A, Hodkinson I. Real-world implementation of video outpatient consultations at macro, meso, and micro levels: mixed-method study. Journal of medical Internet research. 2018 Apr 17;20(4):e9897. doi: 10.2196/jmir.9897.

[3] Shaw SE, Wherton J, Vijayaraghavan S, Morris J, Bhattacharya S, Hanson P, Campbell-Richards D, Ramoutar S, Collard A, Hodkinson I, Greenhalgh T. Advantages and limitations of virtual online consultations in a NHS acute trust: the VOCAL mixed-methods study. Health services and delivery research. 2018 Jul 1;6(21). DOI: 10.3310/hsdr06210

[4] Wherton J, Shaw S, Papoutsi C, Seuren L, Greenhalgh T. Guidance on the introduction and use of video consultations during COVID-19: important lessons from qualitative research. BMJ Leader. 2020 May 17:leader-2020. http://dx.doi.org/10.1136/leader-2020-000262

[5] Peek N, Sujan M, Scott P. Digital health and care in pandemic times: impact of COVID-19. BMJ health & care informatics. 2020;27(1). http://dx.doi.org/10.1136/bmjhci-2020-100166

[6] Steimer M, Leabo J, Wang H, Heyer D, Addison N, Bowles N, Cannon TL, Cuevo R, Ershler WB, Shafer D, Jang S. Remote home monitoring of patients with cancer during the COVID pandemic: A pilot study. JCO Oncology Practice. 2021 Apr:OP-20.

[7] Greenhalgh T, A’Court C. & Shaw S. Understanding heart failure; explaining telehealth – a hermeneutic systematic review. BMC Cardiovasc Disord 2017. 17, 156. https://doi.org/10.1186/s12872-017-0594-2

[8] Powell, J., Gunn, L. E. E., Lowe, P. A. M., Sheehan, B., Griffiths, F., & Clarke, A. (2010). New networked technologies and carers of people with dementia: An interview study. Ageing and Society, 30(6), 1073.

[9] Pols J. Care at a distance: on the closeness of technology. Amsterdam University Press; 2012.

[10] May CR, Eton DT, Boehmer K, Gallacher K, Hunt K, MacDonald S, Mair FS, May CM, Montori VM, Richardson A, Rogers AE. Rethinking the patient: using Burden of Treatment Theory to understand the changing dynamics of illness. BMC health services research. 2014 Dec;14(1):1–1. https://doi.org/10.1186/1472-6963-14-281

[11] Vallo Hult H, Hansson A, Svensson L, Gellerstedt M. Flipped healthcare for better or worse. Health informatics journal. 2019 Sep;25(3):587–97. https://doi.org/10.1177/1460458219833099

[12] Greene J, Hibbard JH. Why does patient activation matter? An examination of the relationships between patient activation and health-related outcomes. Journal of general internal medicine. 2012 May 1;27(5):520–6. doi: 10.1007/s11606-011-1931-2

[13] Barlow J, Wright C, Sheasby J, Turner A, Hainsworth J. Self-management approaches for people with chronic conditions: a review. Patient education and counseling. 2002 Oct 1;48(2):177–87. doi: 10.1016/s0738-3991(02)00032-0

[14] Lorig KR, Sobel DS, Stewart AL, Brown Jr BW, Bandura A, Ritter P, Gonzalez VM, Laurent DD, Holman HR. Evidence suggesting that a chronic disease self-management program can improve health status while reducing hospitalization: a randomized trial. Medical care. 1999 Jan 1:5–14. DOI: 10.1097/00005650-199901000-00003

[15] Donaldson L. Expert patients usher in a new era of opportunity for the NHS: The expert patient programme will improve the length and quality of lives. Bmj. 2003 Jun 12;326(7402):1279–80. doi: 10.1136/bmj.326.7402.1279

[16] Ruckenstein M, Schüll ND. The datafication of health. Annual Review of Anthropology. 2017 Oct 23;46:261–78. https://doi.org/10.1146/annurev-anthro-102116-041244

[17] Ong BN, Rogers A, Kennedy A, Bower P, Sanders T, Morden A, Cheraghi-Sohi S, Richardson JC, Stevenson F. Behaviour change and social blinkers? The role of sociology in trials of self-management behaviour in chronic conditions. Sociology of health & illness. 2014 Feb;36(2):226–38. DOI: 10.1111/1467-9566.12113

[18] Tran VT, Barnes C, Montori VM, Falissard B, Ravaud P. Taxonomy of the burden of treatment: a multi-country web-based qualitative study of patients with chronic conditions. BMC medicine. 2015 Dec;13(1):1–5. https://doi.org/10.1186/s12916-015-0356-x

[19] Sadler E, Wolfe CD, McKevitt C. Lay and health care professional understandings of self-management: a systematic review and narrative synthesis. SAGE open medicine. 2014 Aug 28;2:2050312114544493. doi: 10.1177/2050312114544493

[20] Bury M, Newbould J, Taylor D. A rapid review of the current state of knowledge regarding lay-led self-management of chronic illness: evidence review. London: National Institute for Health and Clinical Excellence; 2005 Dec.

[21] Greenhalgh T. Patient and public involvement in chronic illness: beyond the expert patient. Bmj. 2009 Feb 17;338. doi: https://doi.org/10.1136/bmj.b49

[22] Foster G, Taylor SJ, Eldridge S, Ramsay J, Griffiths CJ. Self-management education programmes by lay leaders for people with chronic conditions. Cochrane database of systematic reviews. 2007(4). doi: 10.1002/14651858.CD005108.pub2

[23] Griffiths C, Foster G, Ramsay J, Eldridge S, Taylor S. How effective are expert patient (lay led) education programmes for chronic disease?. Bmj. 2007 Jun 14;334(7606):1254–6. doi: 10.1136/bmj.39227.698785.47

[24] Peretz D, Arnaert A, Ponzoni NN. Determining the cost of implementing and operating a remote patient monitoring programme for the elderly with chronic conditions: A systematic review of economic evaluations. Journal of telemedicine and telecare. 2018 Jan;24(1):13–21. doi: 10.1177/1357633X16669239.

[25] Castelyn G, Laranjo L, Schreier G, Gallego B. Predictive performance and impact of algorithms in remote monitoring of chronic conditions: a systematic review and meta-analysis. International Journal of Medical Informatics. 2021 Oct 16:104620. DOI: 10.1016/j.ijmedinf.2021.104620

[26] Jonker LT, Plas M, de Bock GH, Buskens E, van Leeuwen BL, Lahr MM. Remote home monitoring of older surgical cancer patients: Perspective on study implementation and feasibility. Annals of surgical oncology. 2021 Jan;28(1):67–78. https://doi.org/10.1245/s10434-020-08705-1

[27] National Health Service. Novel coronavirus (COVID-19) standard operating procedure: COVID Oximetry @home. V1.2. 2020. Accessed from: https://www.england.nhs.uk/coronavirus/wp-content/uploads/sites/52/2020/11/C1396-sop-covid-oximetry-@home-v2-september-21.pdf

[28] National Health Service. Novel coronavirus (COVID-19) standard operating procedure: COVID Virtual Ward. V1.0. 2021. Accessed from: https://www.england.nhs.uk/coronavirus/wp-content/uploads/sites/52/2021/01/C1042-sop-discharge-covid-virtual-ward-13-jan-21.pdf

[29] Vindrola-Padros C, Singh KE, Sidhu MS, Georghiou T, Sherlaw-Johnson C, Tomini SM, Inada-Kim M, Kirkham K, Streetly A, Cohen N, Fulop NJ. Remote home monitoring (virtual wards) for confirmed or suspected COVID-19 patients: a rapid systematic review. EClinicalMedicine. 2021 Jul 1;37:100965. DOI:https://doi.org/10.1016/j.eclinm.2021.100965

[30] Vindrola-Padros C, Sidhu MS, Georghiou T, Sherlaw-Johnson C, Singh KE, Tomini SM, Ellins J, Morris S, Fulop NJ. The implementation of remote home monitoring models during the COVID-19 pandemic in England. EClinicalMedicine. 2021 Apr 1;34:100799. DOI: 10.1016/j.eclinm.2021.100799

[31] Mair FS, Montori VM, May CR. Digital transformation could increase the burden of treatment on patients. British Medical Journal. 2021. 375,n2909. doi: 10.1136/bmj.n2909

[32] Mair FS, May CR. Thinking about the burden of treatment. 2014. 349; g6680. doi: https://doi.org/10.1136/bmj.g6680

[33] Greenhalgh T, Procter R, Wherton J, Sugarhood P, Hinder S, Rouncefield M. What is quality in assisted living technology? The ARCHIE framework for effective telehealth and telecare services. BMC medicine. 2015 Dec;13(1):1–5.https://doi.org/10.1186/s12916-015-0279-6

[34] Borrelli B. The assessment, monitoring, and enhancement of treatment fidelity in public health clinical trials. Journal of public health dentistry. 2011 Jan;71:S52–63. doi: 10.1111/j.1752-7325.2011.00233.x

[35] Walton H, Spector A, Tombor I, Michie S. Measures of fidelity of delivery of, and engagement with, complex, face-to-face health behaviour change interventions: A systematic review of measure quality. British journal of health psychology. 2017 Nov;22(4):872–903. DOI: 10.1111/bjhp.12260

[36] Greenhalgh T, Knight M, Inda-Kim M, Fulop NJ, Leach J, Vindrola-Padros C. Remote management of covid-19 using home pulse oximetry and virtual ward support. bmj. 2021 Mar 25;372. doi: https://doi.org/10.1136/bmj.n677

[37] Alaa, A, Qian, Z., Rashbass, J, Benger, J, van der Scaar, M. (2020). Retrospective cohort study of admission timing and mortality following COVID-19 infection in England. BMJ Open. 2020;10:e042712. doi:10.1136/bmjopen-2020-042712

[38] Mansab F, Donnelly H, Albrecht Kussner JN, Bhatti S, Goyal DK. Oxygen and mortality in COVID-19 pneumonia: a comparative analysis of supplemental oxygen policies and health outcomes across 26 countries. Frontiers in Public Health. 2021;9. https://doi.org/10.3389/fpubh.2021.580585

[39] Fulop et al. A mixed methods evaluation of remote home monitoring models during the COVID-19 pandemic in England (Phase Two evaluation) -Protocol. 2021. Accessed [15/11/2021] from: https://fundingawards.nihr.ac.uk/award/NIHR135016

[40] Lehoux, P., Blume, S. (2000). Technology assessment and the sociopolitics of health technologies. Journal of health politics, policy and law, 25(6), 1083–1120. doi: 10.1215/03616878-25-6-1083

[41] Michie S, Van Stralen MM, West R. The behaviour change wheel: a new method for characterising and designing behaviour change interventions. Implementation science. 2011 Dec;6(1):1–2.

[42] Lyratzopoulos G, Elliott M, Barbiere JM, Henderson A, Staetsky L, Paddison C, Campbell J, Roland M. Understanding ethnic and other socio-demographic differences in patient experience of primary care: evidence from the English General Practice Patient Survey. BMJ quality & safety. 2012 Jan 1;21(1):21–9. http://dx.doi.org/10.1136/bmjqs-2011-000088

[43] Pianori D, Maietti E, Lenzi J, Quargnolo M, Guicciardi S, Adja KY, Fantini MP, Toth F. Sociodemographic and health service organizational factors associated with the choice of the private versus public sector for specialty visits: Evidence from a national survey in Italy. PloS one. 2020 May 7;15(5):e0232827. https://doi.org/10.1371/journal.pone.0232827

[44] English Longitudinal Study of Ageing (ELSA). The data we collect. 2019. Accessed [15/11/2021] from: https://www.elsa-project.ac.uk/the-data-we-collect

[45] Wenz A, Al Baghal T, Gaia A. Language proficiency among respondents: Implications for data quality in a longitudinal face-to-face survey. Journal of survey statistics and methodology. 2021 Feb;9(1):73–93. https://doi.org/10.1093/jssam/smz045

[46] Office for National Statistics. Ethnic group, national identity and religion. N.d. Accessed [15/11/2021] from: https://www.ons.gov.uk/methodology/classificationsandstandards/measuringequality/ethnicgroupnationalidentityandreligion

[47] UK government. Age groups. 2018. Accessed [15/11/2021] from: https://www.ethnicity-facts-figures.service.gov.uk/uk-population-by-ethnicity/demographics/age-groups/latest

[48] Vindrola-Padros C, Chisnall G, Cooper S, et al. Carrying Out Rapid Qualitative Research During a Pandemic: Emerging Lessons From COVID-19. Qualitative Health Research. 2020;30(14):2192–2204. doi:10.1177/1049732320951526

[49] COVID Oximetry @home evaluation interpretation of findings. November 2021. Accessed from: https://www.nuffieldtrust.org.uk/files/2021-11/co-h-interpretation-of-evaluation-findings-final-slide-deck-nov-2021.pdf

[50] Coulter A, Locock L, Ziebland S, Calabrese J. Collecting data on patient experience is not enough: they must be used to improve care. Bmj. 2014 Mar 27;348. doi: https://doi.org/10.1136/bmj.g2225

[51] Mira JJ, Tomás O, Virtudes-Pérez M, Nebot C, Rodríguez-Marín J. Predictors of patient satisfaction in surgery. Surgery. 2009 May 1;145(5):536–41. doi: 10.1016/j.surg.2009.01.012.

[52] Vassilev I, Rogers A, Sanders C, Kennedy A, Blickem C, Protheroe J, Bower P, Kirk S, Chew-Graham C, Morris R. Social networks, social capital and chronic illness self-management: a realist review. Chronic illness. 2011 Mar;7(1):60–86. https://doi.org/10.1177/1742395310383338

[53] Russell S, Ogunbayo OJ, Newham JJ, Heslop-Marshall K, Netts P, Hanratty B, Beyer F, Kaner E. Qualitative systematic review of barriers and facilitators to self-management of chronic obstructive pulmonary disease: views of patients and healthcare professionals. NPJ primary care respiratory medicine. 2018 Jan 17;28(1):1–3. https://doi.org/10.1038/s41533-017-0069-z

[54] Walton H, Spector A, Roberts A, Williamson M, Bhatt J, Tombor I, Michie S. Developing strategies to improve fidelity of delivery of, and engagement with, a complex intervention to improve independence in dementia: a mixed methods study. BMC medical research methodology. 2020 Dec;20(1):1–9. https://doi.org/10.1186/s12874-020-01006-x

[55] Boulton ER, Horne M, Todd C. Multiple influences on participating in physical activity in older age: Developing a social ecological approach. Health Expectations. 2018 Feb;21(1):239–48. DOI: 10.1111/hex.12608

[56] Roberts SH, Bailey JE. Incentives and barriers to lifestyle interventions for people with severe mental illness: a narrative synthesis of quantitative, qualitative and mixed methods studies. Journal of advanced nursing. 2011 Apr;67(4):690–708. DOI: 10.1111/j.1365-2648.2010.05546.x

