## Supplementary files 1-3 for "Patients’ and carers’ experiences of, and engagement with remote home monitoring services for COVID-19 patients: a rapid mixed-methods study"

**Appendix 1. Detailed study methodology for patient and carer survey and interviews**

### i) NATIONAL STUDY OF PATIENT EXPERIENCES

The aim of the national study of patient experiences was to analyse patient experiences of care in sites across England.

This aspect of the study included data from national patient or carer surveys.

### National surveys (patients)

#### Sample and recruitment

##### Selection of sites

Twenty-eight services were included in our national evaluation. Each site had a research lead (MS, CVP, HW, JB, IL, LH) to support data collection and act as an ongoing point of contact for the site.

To obtain maximum variation, we sampled services based on a range of criteria, including the setting (primary care or secondary care), type of model (pre-hospital, early discharge, both), mechanism for patient monitoring (paper-based, app, both), geographic location (across different areas of the country), timing of implementation (implemented since wave 1 of the pandemic or recently implemented) and involvement in the evaluation with the other evaluation partners (Imperial and IAU).

Sites were recruited through an expression of interest process whereby we presented our study at local and national meetings and asked sites to express interest in participating. Clinical Research Networks facilitated the setup of sites and local governance approvals. Some sites were identified through our phase 1 evaluation.

##### Patient survey

25/28 sites agreed to conduct the patient and carer survey. To participate in our survey, participants needed to be:

I. 18 or over,
II. Proficient in English (or one of the following languages: Polish, Bengali, Urdu, Punjabi, French and Portuguese),
III. Eligible to receive COVI-19 remote home monitoring services, and must also have been offered and received COVID-19 remote home monitoring.

National and local eligibility for COVID-19 remote home monitoring varies. We were flexible within our sampling to take into account both national and local eligibility criteria. For reference, the national eligibility guidelines are as follows: To be eligible for receiving COVID-19 remote home monitoring patients must have a confirmed or suspected diagnosis of COVID-19 plus be one of the following: (a) Symptomatic with COVID-19 & aged 65 years or older, b) Symptomatic with COVID-19 & under 65 years but ‘clinically extremely vulnerable’ (using the Clinically extremely vulnerable to COVID list) to COVID (CO@h Standard operating procedure, 2020).

For the patient survey, NHS staff from participating services sent the patient survey to patients (or their carers if applicable) onboarded onto the service between 1^st^ January 2021-11^th^ June 2021. NHS sites decided how to best disseminate the survey to their patients (either via post or text/email). All survey sites were asked to keep a record of the number of surveys they have sent out to determine patient response rates.

If patients were not able/willing to take part in the survey, they were given the option to ask their carer or family member to complete the survey on their behalf, reflecting on the patient’s experience with the service. The survey was sent to patients who have received care at participating sites by NHS staff. Patients/carers returned completed surveys directly to the study team for analysis, either electronically through REDCap or via post using pre-paid envelopes. In addition to English, we also offered participants the opportunity to receive an information sheet and survey in six other languages (Polish, Bengali, Urdu, Punjabi, French and Portuguese).

#### Measures

##### Patient survey

We developed patient surveys specifically for this study.

The aim of the patient survey was to capture the experiences of patients who received COVID-19 remote home monitoring, and their engagement with the COVID-19 remote home monitoring service. We developed a patient experience survey for this purpose. The survey included closed questions focused on: the service that patients have received, their experience with the service and their engagement with the service. As part of the survey, we also asked questions about patients experience of analogue vs tech-enabled models. These questions were followed by a single open text question at the end to give participants the opportunity to share any wider thoughts. Survey and interview questions were informed by relevant service documentation (NHS, 2020, NHS 2021), theoretical frameworks relating to social, political and technical contexts (Greenhalgh, 2017; Powell, 2010; Greenhalgh, 2015; Lehoux & Blume, 2000) and behaviour (Michie et al, 2011), and previous literature on engagement [Borrelli, 2011; Walton et al, 2017]. We also included a section at the end of the survey to ask about participants’ socio-demographic characteristics (including questions on gender, age, ethnicity, education, employment, disability, sexuality, first language and geographical region). Questions on demographic characteristics were informed by previous literature (Lyratzopoulos, 2012; Pianori, 2020; ELSA, 2019; Wenz 2021; ONS, UK government, 2018). To reduce burden and maximise response rates, the patient survey was designed to take between 15 and 30 minutes to complete. The survey was delivered using the online platform REDCap.

Survey questions were reviewed, and sense-checked by our clinical advisory group, PPI group and our cohort of 70@70 nurses. The theoretical frameworks were used as a sensitising device to inform the development of questions in the surveys and interviews. The patient survey was piloted with our PPI group and some members of the public. In response to feedback for the patient survey, and to make the survey more accessible, we amended some of the questions, increased the font size, reduced the number of questions and added definitions for key terms (such as oximeter).

#### Data collection

##### Patient survey

For the patient survey, patients were approached by NHS staff to take part in a survey in one of two different ways: 1) if the patient was monitored through the use of an app, they received an SMS/email with a link to the online survey, 2) if the patient was monitored through regular phone calls and a paper-based recording method, they received the survey in the post (with a pre-paid addressed envelope). Whilst most surveys were distributed at discharge, some sites chose to distribute the paper survey at onboarding and then remind patients at discharge to complete the survey. NHS staff distributed the online and paper version of the survey so the research team had no access to patient information. Both survey options (online and paper) included prefacing information with a background to the study, potential risks, indicating voluntary participation, anonymity and a description of how the data will be used. This page also included boxes that patients/carers were asked to tick to indicate their consent to take part in the study. The method of administering the survey to patients (i.e. NHS staff sending to patients) meant that there were no reminders. Due to research capacity, we were unable to conduct the survey with patients over the phone. Data collection took place between March and June 2021.

#### Data management

Surveys were returned to the research team, either electronically through REDCap (patient surveys), or by posting completed surveys in pre-paid envelopes to our RSET team members at the Nuffield Trust or UCL (patient surveys only). Surveys received via post were stored securely in locked filing cabinets within secure Nuffield Trust or UCL offices. Data from patient surveys sent via post were inputted into REDCap by members of the research team. Data from the patient surveys were directly stored in the UCL Data Safe Haven via REDCap, as this will include identifiable information (e.g. postcode data). Data from the completed surveys were stored securely using password protected spreadsheets to which only the RSET and BRACE researchers had access to.

#### Analysis

Sites were characterised with respect to their population size, the proportion in urban versus rural areas, and the proportion in the most and least deprived areas (with respect to national quintiles). For sites based on CCG areas we calculated these characteristics using publicly available data at lower super output area (LSOA) level mapped to CCGs, while for trust-based sites we used data derived from inpatient Hospital episode statistics (HES) admissions during the financial year 2019/20, in addition to web searches for the trust catchment populations.

The quantitative survey data were analysed using SPSS statistical software (version 25). Descriptive statistics, multivariate and univariate analyses were conducted to compare patient experiences of the service across patient groups and service models (as reported by patients and carers). We offered to carry out site-specific analyses of patient experience data for participating sites.

### ii) CASE STUDIES OF PATIENT EXPERIENCES

The aim was to document in-depth patient experiences of care in a sample of 17 sites. 15/17 sites took part in the patient survey.

This aspect of the study included data from interviews with patients/carers.

#### Sample and recruitment

##### Site selection

A smaller sample of the overall study sites were included as case studies in order to conduct a more in-depth analysis of implementation, patient and staff experiences.

Seventeen of the twenty-eight sites were selected as in-depth case study sites using the aforementioned criteria (see national site selection). Four of the 17 sites were purposively selected by NHSX for a more in-depth analysis of patient experiences of tech-enabled models of care; sites using different tech-enabled platform were selected.

##### Patient interviews

We aimed to interview up to six participants (patients or their carer) who had received, disengaged or declined with COVID-19 remote home monitoring from each site. To participate in our patient or carer interviews, participants needed to be 18 or over, proficient in English (or one of the following languages: Polish, Bengali, Urdu, Punjabi, French and Portuguese), eligible to receive COVID-19 remote home monitoring services, and must also have been offered and either received or refused the service. If patients were not able/willing to take part in the interview, patients were asked by site coordinators if their carer (if they have one) could be approached to capture their perceptions of the patient’s journey and overall experience with the service.

We asked the main contact person at each site (the study coordinator), to identify a convenience sample of 4-6 patients (or their carers). To identify potential participants, the study coordinator contacted potential participants to see if they were happy to be approached by a researcher. If they agreed, the researcher contacted the patient or their carer via telephone or email to discuss the study. NHS staff identified patients on behalf of the study team using a purposive sampling approach. To be inclusive and capture a wide range of views, we asked sites to select patients with different characteristics (e.g. age, gender, ethnicity, deprivation score (by postcode), employment status, and comorbidities). We also asked sites to identify interviewees who had declined or disengaged from the service and those who used different data submission methods (if applicable).

If the patient or their carer was contacted via phone, they were asked if a participant information sheet and consent form can be sent via email. If they preferred post, both of these documents were sent via post with a pre-paid addressed envelope so they could return the signed consent form to the team. If the patient was contacted via email, the participant information sheet and consent form were sent in a subsequent email and the patient was given the option to schedule a call with the researcher to discuss the study. The participant information sheet contained information on the study, potential risks and a description of how the data will be used to ensure informed and voluntary participation. If the patient or their carer agreed to take part in the study, they were asked to email back the signed consent form (scanned forms or typewritten/electronic signature). If patients were not able/willing to take part in the interview, we asked patients if we can approach their carer (if they have one) to capture their perceptions of the patient’s journey and overall experience with the service.

#### Measures

##### Patient and carer interviews

Patient and carer interview topic guides included questions for patients who had received COVID-19 remote home monitoring services, those who had declined to receive the service and those who disengaged from the service. The interviews with patients and carers focused on documenting their journeys of remote home monitoring, their experiences of being ill and monitored at home, experiences with escalation and discharge, their engagement with the service, and recommendations for improving these models. Interview questions (as with survey questions) were informed by relevant service documentation (NHS, 2020, NHS 2021) and literature (Greenhalgh, 2017; Powell, 2010; Greenhalgh, 2015; Lehoux & Blume, 2000; Michie et al, 2011; Borrelli, 2011; Walton et al, 2017).

During the interview, we asked patients/carers some brief questions relating to socio-demographic characteristics including whether they are a patient or carer, age, gender, ethnicity, how many people they live with, education and qualifications, employment status, English as a first language, disability and postcode (the latter to be used as indicator of social deprivation) (Lyratzopoulos, 2012; Pianori, 2020; ELSA, 2019; Wenz 2021; ONS, UK government, 2018). We emphasised that as with all parts of the interview, these questions are optional.

We intended to conduct ‘think alouds’ with patients who had used tech-enabled data submission from the four sites selected for in-depth analysis of tech-enabled platforms; however, patients did not have access to the platforms after discharge and recall of the use of these platforms during their illness was poor. Consequently, think aloud methodology was discontinued for patient interviews.

To determine whether questions were appropriate and relevant, we discussed the interview topic guides with our PPI members and the 70@70 nurses. The topic guides were amended accordingly.

#### Data collection

The researcher arranged a time to carry out the interview. Each site had a different lead researcher who conducted the interviews and liaised with sites on an on-going basis. Interviews were conducted by six researchers (MS, CV, HW, LH, IL, JB). Interviews were carried out via telephone or an online platform (e.g. Zoom or MS Teams) as preferred by the participant. Data collection for interviews was conducted between February and June 2021.

#### Data management

All interviews were semi-structured, audio recorded (subject to consent being given), transcribed verbatim by a professional transcription service (TP Transcription limited), anonymised and kept in compliance with the General Data Protection Regulation (GDPR) 2018 and Data Protection Act 2018.

#### Analysis

For patient interviews, data collection and analysis was carried out in parallel and facilitated through the use of RAP sheets as explained in Vindrola-Padros et al. (2020). RAP sheets were developed per site to facilitate cross-case comparisons and per population (to make comparisons between sub-groups). The categories used in the RAP sheets were based on the questions included in the interview topic guide, maintaining flexibility to add categories as the study is ongoing.

**Appendix 2. Site characteristics**

| **Characteristic** | | **Number of sites** |
| --- | --- | --- |
| Region | London  South West  South East  North West  North East  East Midlands  East of England  Yorkshire and Humber | 5  7  5  5  2  2  1  1 |
| Size of population | <250,000  250,000-500,000  500,000-1 million  >1 million | 4  8  11  5 |
| % Urban (% rural) | 65-80 (20-35)  80-95 (5-20)  95-100 (0-5) | 8  10  10 |
| Deprivation | **% of population in most deprived quintile**  0-15  15-25  25-50  50+  **% of population in least deprived quintile**  0-15  15-25  25-50  50+ | 13  7  7  1  13  7  8 |
| Ethnicity (% of population non-white) | 0-5  5-15  15-30  30-50  50-65 | 9  10  4  3   2 |

**Appendix 3. Interview topic guide**

**PATIENTS**

**INTRODUCTION**

The interview should last between 30 and 60 mins, depending on how much you would like to say. We will ask questions about how you found the experience of monitoring and recording your COVID symptoms at home, and any further advice you received – we call this “COVID care at home”. We will feedback the results of this evaluation to local and national NHS and public health services, and results will be made available to the general public too.

If you do not want to answer a question, you do not have to, and if you feel uncomfortable or tired we can stop the interview at any point. Let us know if you’d like a break or to come back later. We can also carry out the interview in two halves if that is easier for you to manage. Have you got any questions before we start?

**Questions for those who have received CO@h**

| **Main question** | **Follow up questions (prompts)** |
| --- | --- |
| 1. Please tell me a bit about yourself, | - Do you live by yourself or with others? - How long have you lived in your neighbourhood? - Do you have any family or friends living close by? - Is English your first language? If N, ask what is their first language. |
| **FINDING OUT ABOUT THE SERVICE**   2. Can you tell me the story of how you ended up being referred to COVID care at home? | a. How were you referred to the service? b. Who did you speak to and when? (virtual/face to face? How were you involved in the assessment process?) *(for assessment/triage)* c. When you were first told about the service, how was it described to you? d. What were your first impressions of COVID care at home? *(prompt about whether they found it reassuring or not)*    a. Positive impressions?    b. Did you have any concerns/worries? |
| **DESCRIBING COVID CARE AT HOME**  3. COVID care at home is carried out slightly differently in different parts of the country. Please can you tell me about what receiving COVID care at home has involved for you? | a. What equipment were you given?  b. How/when was the pulse oximeter delivered to you? Can you tell me how it works?  c. What symptoms did you have to monitor? For each, ask how often.  d. How did you have to record your symptoms? (e.g. paper, digital app, telephone line)  e. Were you offered a choice in how you recorded your symptoms?  f. Who did you speak to? (& how often – monitoring)  g. Have family members/carers been involved? If so, how? (if relevant)  h. Overall, how did you feel about recording and monitoring your symptoms? *(prompt about whether they found it reassuring or not)* |
| **INFORMATION RECEIVED ON COVID CARE AT HOME AT ONBOARDING**  4 a. Did you receive information on COVID care at home **in person** from a member of the care team?  If no, move to 4b.  4b. Did you receive information on COVID care at home **over the telephone or a video call** from a member of the care team?  If no, move to 4c.  4c. Were you directed to any information **to read or watch on a website** on COVID care at home?  If no, move to 4d.  4d. Were you directed to any information **to read or watch on an app** on COVID care at home?  If no, move to 5. | a. i) If yes, what information did you receive? (*Monitoring symptoms? Using oximeter? Recording symptoms? Seeking further advice?)*  ii) Who gave you the information?  Was it in your first language?  iii) How easy was it to understand the information?  iv) Did the person you spoke to describe the readings and what they mean in relation to your everyday symptoms and experience?  b. i) If yes, what information did you receive? (*Monitoring symptoms? Using oximeter? Recording symptoms? Seeking further advice?)*  ii) Who gave you the information?  iii) Was it in your first language?  iv) How easy was it to understand the information?  c. i) If yes, were you able to access the information you needed on the website?  ii) If no, why not?  If yes, what information did you receive? (*Monitoring symptoms? Using oximeter? Recording symptoms? Seeking further advice*?)  iii) Was it in your first language?  iv) How easy was it to understand the information?  d. i) If yes, were you able to access the information you needed on the app?  If no, why not?  If i) is yes, what information did you receive? *(Monitoring symptoms? Using oximeter? Recording symptoms? Seeking further advice?)*  iii) Was it in your first language?  iv) How easy was it to understand the information? |
| **CARRYING OUT THE MONITORING**  **OXIMETER**  5. How did you find using the oximeter to monitor your oxygen levels? *(may like to prompt about how often they used it)*  **OTHER SYMPTOMS**  6. Overall, how did you find monitoring your other symptoms (pulse heart rate/temperature/symptoms) at home? *(may like to prompt about how often they monitored other symptoms)* | a. What helped you to use it?  b. What got in the way?  c. What did you do when you had problems? (prompt about type of support and usefulness)  What could be changed to make it easier to use the oximeter?  a. What worked well?  b. What got in the way?  c. What did you do when you had problems? (prompt about type of support and usefulness)  d. What could be changed to make it easier to monitor your other symptoms?  e. Was there any parts of monitoring that you were uncertain about?  f. Did you seek further advice from anyone about monitoring your symptoms? Prompt – who, when, how? |
| **CARRYING OUT RECORDING**  7. Overall, how did you find recording your readings (blood oxygen levels/symptoms/pulse heart rate/temperature)? *(using an app / diary / both) (prompt about how often they recorded their readings)* | a. What worked well?  b. What got in the way?  c. What did you do when you had problems? (prompt about type of support and usefulness)  d. What could be changed to make it easier to monitor your other symptoms?  e. Was there any part of recording your symptoms that you were uncertain about? *(prompt about whether they felt confident)*  r. Did you seek further advice from anyone about recording? Prompt – who, when, how? |
| **COMMUNICATING READINGS TO MEMBER OF THE TEAM**  **8. How have you found sending or communicating your symptoms and readings to the COVID care at home team? (if needed)** | a. What worked well?  b. What got in the way?  c. What did you do when you had problems? (prompt about type of support and usefulness)  d. What could be changed to make it easier to communicate your readings to the COVID care at home team?  e. Was there any part of communicating your readings to a member of the team that you were uncertain about?  r. Did you seek further advice from anyone about communicating your readings? Prompt – who, when, how? |
| **SEEKING FURTHER ADVICE**  8. Have you had to seek further support and help (escalate your care) because of the readings given by your oximeter or because of other things such as a change in symptoms? | a. How did this go?  b. What did this involve?  c. What was your experience of being sent for further support and help (e.g. escalated or admitted to hospital)?  d. What were you instructed to do? (i.e. dial 111, dial 999, go to A&E)?  e. Did you self-escalate your care if necessary? Why or why not? *(prompt on how they made the decision that they needed to seek further help)*  f. What helped you to seek further support?  g. What got in the way of seeking further support?  h. What could be changed to make it easier? |
| **DISCHARGE**  9. What was your understanding about what would happen once you are discharged from COVID care at home? | a. How did you feel after being discharged from COVID care at home? (prompt about whether they found it reassuring or not?) b. Since being discharged from COVID care at home, what other services/support have you accessed? *(If discharged from COVID care at home).*    i. What were these?    ii. How often have you had to access these? |
| **RECOMMENDATIONS**  10. If a friend who was in a similar position to you at the start of your illness was offered COVID care at home, would you recommend it to them  a) over hospital care?  b) over no monitoring, with the option to access usual services as needed.  11. Do you have any recommendations to improve the service? | A) If yes, why? If no, why not?  B) If yes, why? If no, why not?  If yes, what? |
| 12. Is there anything else that you would like to say about what we have talked about? |  |

**If participants did not want to receive COVID care at home**

Could ask Q1-5, and then:

| **Main question** | **Follow up questions (prompts)** |
| --- | --- |
| 1. Please tell me a bit about yourself | - Do you live by yourself or with others? - How long have you lived in your neighbourhood? - Do you have any family or friends living close by? - Is English your first language? If N, ask what is their first language. |
| **FINDING OUT ABOUT THE SERVICE**   2. Can you tell me the story of how you ended up being referred to COVID care at home? | a. How were you referred to the service? b. Who did you speak to and when? (virtual/face to face? How were you involved in the assessment process?) *(for assessment/triage)* c. When you were first told about the service, how was it described to you? d. What were your first impressions of COVID care at home? *(prompt about whether they found it reassuring or not)*    a. Positive impressions? e. Did you have any concerns/worries? |
| **EXPECTATIONS OF COVID CARE AT HOME**   3. Please tell me about what you thought receiving COVID care at home would involve | a. What equipment would you be given?  c. What symptoms would you have had to monitor? For each, ask how often.  d. How would you have had to record your symptoms? (e.g. paper, digital app, telephone line)  e. Were you offered a choice in how you could have recorded your symptoms?  f. Who did you speak to?  h. Overall, how did you feel about the prospect of recording and monitoring your symptoms? *(prompt about whether they found it reassuring or not)* |
| **INFORMATION RECEIVED ON COVID CARE AT HOME AT ONBOARDING**  4 a. Did you receive information on COVID care at home **in person** from a member of the care team?  If no, move to 4b.  4b. Did you receive information on COVID care at home **over the telephone or a video call** from a member of the care team?  If no, move to 4c.  4c. Were you directed to any information **to read or watch on a website** on COVID care at home?  If no, move to 4d.  4d. Were you directed to any information **to read or watch on an app** on COVID care at home?   4. If no, move to 5. | a. i) If yes, what information did you receive? (*Monitoring symptoms? Using oximeter? Recording symptoms? Seeking further advice?)*  ii) Who gave you the information?  Was it in your first language?  iii) How easy was it to understand the information?  iv) Did the person you spoke to describe the readings and what they mean in relation to your everyday symptoms and experience?  b. i) If yes, what information did you receive? (*Monitoring symptoms? Using oximeter? Recording symptoms? Seeking further advice?)*  ii) Who gave you the information?  iii) Was it in your first language?  iv) How easy was it to understand the information?  c. i) If yes, were you able to access the information you needed on the website?  ii) If no, why not?  If yes, what information did you receive? (*Monitoring symptoms? Using oximeter? Recording symptoms? Seeking further advice*?)  iii) Was it in your first language?  iv) How easy was it to understand the information?  d. i) If yes, were you able to access the information you needed on the app?  If no, why not?  If i) is yes, what information did you receive? (Monitoring symptoms? Using oximeter? Recording symptoms? Seeking further advice?)  iii) Was it in your first language?  iv) How easy was it to understand the information?   a. |
|  | a. |
| **REASONS FOR DECLINING**   5. Why did you choose not to take part in the CO@h remote monitoring? 6. Could anything be changed to make you want to receive COVID care at home more? | a. b. Did anything get in the way? If so, what? c. How did this get in the way? d. If so, what? e. How would this help? |
| **OTHER CARE/SERVICES ACCESSED**   7. Did you have any other type of monitoring? | f. If so, what? g. How did you find this? |
| 8. Have you had to seek further support and help? (e.g. GP or A&E) | a. If so, what? |
| 9. Is there anything else you’d like to say about what we have talked about? | a. How would this help? |

**If participants withdrew from receiving COVID care at home**

| **Main question** | **Follow up questions (prompts)** |
| --- | --- |
| 1. Please tell me a bit about yourself | - Do you live by yourself or with others? - How long have you lived in your neighbourhood? - Do you have any family or friends living close by? - Is English your first language? If N, ask what is their first language. |
| **FINDING OUT ABOUT THE SERVICE**   2. Can you tell me the story of how you ended up being referred to COVID care at home? | a. How were you referred to the service? b. Who did you speak to and when? (virtual/face to face? How were you involved in the assessment process?) *(for assessment/triage)* c. When you were first told about the service, how was it described to you? d. What were your first impressions of COVID care at home? *(prompt about whether they found it reassuring or not)*    a. Positive impressions? e. Did you have any concerns/worries? |
| **DESCRIBING COVID CARE AT HOME**   3. COVID care at home is carried out slightly differently in different parts of the country. Please can you tell me about what receiving COVID care at home has involved for you? | a. What equipment were you given?  b. How/when was the pulse oximeter delivered to you? Can you tell me how it works?  c. What symptoms did you have to monitor? For each, ask how often.  d. How did you have to record your symptoms? (e.g. paper, digital app, telephone line)  e. Were you offered a choice in how you recorded your symptoms?  f. Who did you speak to? (& how often – monitoring)  g. Have family members/carers been involved? If so, how? (if relevant)  h. Overall, how did you feel about recording and monitoring your symptoms? *(prompt about whether they found it reassuring or not)* |
| **INFORMATION RECEIVED ON COVID CARE AT HOME AT ONBOARDING**  4 a. Did you receive information on COVID care at home **in person** from a member of the care team?  If no, move to 4b.  4b. Did you receive information on COVID care at home **over the telephone or a video call** from a member of the care team?  If no, move to 4c.  4c. Were you directed to any information **to read or watch on a website** on COVID care at home?  If no, move to 4d.  4d. Were you directed to any information **to read or watch on an app** on COVID care at home?  If no, move to 5. | a. i) If yes, what information did you receive? (*Monitoring symptoms? Using oximeter? Recording symptoms? Seeking further advice?)*  ii) Who gave you the information?  Was it in your first language?  iii) How easy was it to understand the information?  iv) Did the person you spoke to describe the readings and what they mean in relation to your everyday symptoms and experience?  b. i) If yes, what information did you receive? (*Monitoring symptoms? Using oximeter? Recording symptoms? Seeking further advice?)*  ii) Who gave you the information?  iii) Was it in your first language?  iv) How easy was it to understand the information?  c. i) If yes, were you able to access the information you needed on the website?  ii) If no, why not?  If yes, what information did you receive? (*Monitoring symptoms? Using oximeter? Recording symptoms? Seeking further advice*?)  iii) Was it in your first language?  iv) How easy was it to understand the information?  d. i) If yes, were you able to access the information you needed on the app?  If no, why not?  If i) is yes, what information did you receive? (Monitoring symptoms? Using oximeter? Recording symptoms? Seeking further advice?)  iii) Was it in your first language?  iv) How easy was it to understand the information?   a. |
| **REASON FOR WITHDRAWING FROM CO@H**   5. Why did you choose to withdraw from receiving COVID care at home? | b. - Did anything get in the way? If so, what?   - How did this get in the way? |
| **CARRYING OUT THE MONITORING (if applicable)**  **OXIMETER**   6. How did you find using the oximeter to monitor your oxygen levels?   **OTHER SYMPTOMS**   7. Overall, how did you find monitoring your other symptoms (pulse heart rate/temperature/symptoms) at home? | a. What helped you to use it?  b. What worked less well?  c. What did you do when you had problems? (prompt about type of support and usefulness)  What could be changed to make it easier to use the oximeter?  a. What worked well?  b. What worked less well  c. What did you do when you had problems? (prompt about type of support and usefulness)  d. What could be changed to make it easier to monitor your other symptoms? |
| **CARRYING OUT RECORDING (If applicable)**   8. Overall, how did you find recording your readings (blood oxygen levels/symptoms/pulse heart rate/temperature)? *(using an app / diary / both)* | a. What worked well?  b. What worked less well?  c. What did you do when you had problems? (prompt about type of support and usefulness)  d. What could be changed to make it easier to monitor your other symptoms?  e. Was there any part of recording your symptoms that you were uncertain about?  r. Did you seek further advice from anyone about recording? Prompt – who, when, how? |
| **COMMUNICATING READINGS TO MEMBER OF THE TEAM (If applicable)**  9. How have you found sending or communicating your symptoms and readings to the person taking the readings (if needed) | a. What worked well?  b. What worked less well?  c. What did you do when you had problems? (prompt about type of support and usefulness)  d. What could be changed to make it easier to communicate your readings to a member of the team?  e. Was there any part of communicating your readings to a member of the team that you were uncertain about?  f. Did you seek further advice from anyone about communicating your readings? Prompt – who, when, how? |
| **SEEKING FURTHER ADVICE (If applicable)**   9. Have you had to seek further support and help (escalate your care) because of the readings given by your oximeter or because of other things such as a change in symptoms? | a. How did this go? b. What did this involve? c. What was your experience of being sent for further support and help (e.g. escalated or admitted to hospital)? d. What were you instructed to do? (i.e. dial 111, dial 999, go to A&E)? e. Did you self-escalate your care if necessary? Why or why not? f. What helped you to seek further support? g. What got in the way of seeking further support? h. What could be changed to make it easier? |
| **OTHER CARE/SERVICES ACCESSED**  10. Did you have any other type of monitoring? | c. - If so, what? d. -How did you find this? |
| 11. Have you had to seek further support and help? (e.g. GP or A&E) | e. - If so, what? |
| **RECOMMENDATIONS**  12. Could anything be changed to make you want to receive COVID care at home more?  13. If a friend who was in a similar position to you at the start of your illness was offered COVID care at home, would you recommend it to them  a) over hospital care?  b) over no monitoring, with the option to access usual services as needed.  14. Do you have any recommendations to improve the service? | a. If so, what? b. How would this help?   A) If yes, why? If no, why not?  B) If yes, why? If no, why not?  If yes, what? |
| 15. Is there anything else you’d like to say about what we have talked about? |  |

**Interview questions on demographic characteristics (asked at the end of the interview)**

- Patient or carer? (relationship with patient, if carer)
- Gender
- Age
- How many people do you live/cohabit with?
- Which of these best describes your living arrangement? *Please select one answer (I own my home outright, I own my home with a mortgage, I rent from local authority/housing association, I rent privately, Other (e.g. living with family/friends), prefer not to say)*
- Ethnicity of patient (and family member if applicable)
- At what age did you complete your continuous full time education? *(__ years/never went to school, do not wish to answer)*
- Which of these best describes your highest *educational* qualification? *(Please select one answer) (No formal qualification, GCSE/CSE/O level or equivalent, A level/AS level or equivalent, Degree level or higher, Other (please specify), do not wish to answer)*
- Which of these best describes your current work situation? *(please tick all that apply) (Working full time, working part time, self-employed, student in higher education, unemployed, homemaker, retired, furloughed under COVID-19, Full time carer (of dependent child or adult), not in work due to poor health or disability,, Other (if other please describe), do not wish to answer).*
- Is English your first language? *(yes/no/do not wish to answer)*
- Prior to your current illness, are your day-to-day activities limited because of a health problem or disability which has lasted, or is expected to last, at least 12 months?? (Includes problems which are due to old age.) (yes, limited a lot/yes, limited a little, no/do not wish to answer)
- What is your Postcode? (optional)
