## Supplementary file 4 for "Patients’ and carers’ experiences of, and engagement with remote home monitoring services for COVID-19 patients: a rapid mixed-methods study"

*Please quote this ID number if you would like to withdraw:*

### COVID CARE AT HOME: PATIENT AND CARER EXPERIENCE SURVEY

#### Background information

Please read the information sheet and this information before completing the survey.

Thank you for taking the time to look at our survey. The responses you provide will help us to understand more about the experiences of patients and carers (such as family members or friends who have provided care) who have received COVID care at home. The survey is part of a larger national study which is looking at the impact of COVID care at home.

The survey may take between 15 and 30 minutes to complete. The questions in the survey will ask you about the service that you (or your family member) received and your experience receiving and engaging with the service, and some questions about you (and your family member - if applicable). Please feel free to complete this survey with your family member/friend if you would like to. Please do not include any identifiable information (e.g. your name) in your responses.

After completing the survey, **please post your survey responses back to us (the research team) using the pre-paid envelope provided**.

The survey is part of a larger study which is funded by the UK National Institute for Health Research (NIHR).

This study has been reviewed and given favourable ethical opinion by the London-Bloomsbury Research Ethics Committee (REC reference: 21/HRA/0155).

The principal investigator for this study is Professor Naomi Fulop. If you have any questions about the survey, please contact Dr Holly Walton by or telephone: 020 3108 3068.

**Important information about your data**
By completing this questionnaire, you are giving your consent for the information you have provided to be used by researchers at University College London, University of Birmingham and RAND Europe for this study.

The information obtained from surveys will be stored securely and managed in accordance with the UK Data Protection Act (2018) and General Data Protection Regulation (GDPR) 2018 and in accordance with the University College London, RAND Europe (a research consultancy company), Nuffield Trust and University of Birmingham's policies for data storage and management. Your personal data will be handled securely and anonymised after analysis and before publication.

You can withdraw your data up until 14 days after the date that you complete the survey. You can withdraw from the survey at any time during completion by exiting the survey. You will not be identifiable and all of your responses will remain anonymous.

For more information about the survey, please read the Participant information sheet (date: 10/02/2021 version 1.2)

Thank you so much for taking time to read this information.

If you would like to complete this survey, please tick all of the statements on Page 2 ('Section A. Consent to take part') and continue to complete the survey.

#### Section A. Consent to take part

By ticking each box below and returning the survey to the research team, you are providing consent to take part in this survey. **Please tick all boxes if you would like to take part** (1-6):

| | Please tick: |
|---|---|
| 1. I have read the information provided for this study (information sheet dated 10/02/2021, version 1.2). I have had the opportunity to consider the information, ask questions and have had these answered satisfactorily. | ☐ |
| 2. I understand that my participation in this survey is voluntary and that I am free to withdraw up to 14 days after completing the survey. I understand if I withdraw, the data I have provided up until that time will be deleted. | ☐ |
| 3. I understand that all personal information will be used for purposes explained to me, will remain confidential and that all efforts will be made to ensure I cannot be identified. | ☐ |
| 4. I understand that any identifiable data will be stored securely and will not be shared with anyone outside of the research team. | ☐ |
| 5. I give permission for the identifiable data I provide to be archived at University College London for up to three years after the end of the project. The anonymised data will be archived for up to 20 years. | ☐ |
| 6. I agree to take part in this survey. | ☐ |

If you have ticked boxes 1-6 above, please proceed to Section B.

#### Section B. Type of respondent

1. **Are you the patient who received COVID care at home or a carer (such as a family member or friend?)** *(please tick)*

☐ Patient or service user

☐ Carer (e.g. family member or friend of someone who has received COVID care at home)

#### Section C. Your experience of COVID care at home

*Please answer the below survey questions about your experience of COVID care at home. The first questions relate to the service you received from COVID care at home.*

**Note for carers completing this survey: If you are answering this survey as a carer, please respond to the questions in this section on behalf of your family member/friend who received COVID care at home (i.e. 'you' refers to you and your family member/friend).*

2. **How were you referred to the COVID care at home service?** *(please tick)*

☐ By a community healthcare professional (e.g. GP or community clinics)

☐ By the hospital (after attending the emergency department)

☐ By the hospital (after being discharged as an inpatient)

☐ Other (please specify): ____________________

☐ Not sure

| | Yes | No | Not sure |
|---|---|---|---|
| 3. **Did you receive an oximeter?** (please tick) *(*An oximeter is a small machine that is placed on your finger. Oximeters are used to measure your blood oxygen levels)* | ☐ | ☐ | ☐ |
| 4. **Were you given information on how to use the oximeter?** *(please tick)* | ☐ | ☐ | ☐ |
| 5. **Were you given information on how to take and record your readings?** (please tick) *(*Readings include oxygen saturation levels, temperature or heart rate)* | ☐ | ☐ | ☐ |
| 6. **Were you given information about what to do if your oxygen levels dropped below the recommended levels?** *(please tick)* | ☐ | ☐ | ☐ |

7. **How was the information provided to you?** *(please tick)*

- ☐ Written information
- ☐ Over the telephone
- ☐ Online information (e.g. video)
- ☐ Other (please specify): ____________
- ☐ Not applicable

| | | | | | | |
|---|---|---|---|---|---|---|
| 8. **How did you find understanding the information about COVID Care at home that you were provided with?** *(please circle one response)* | Very easy | Easy | Neutral | Difficult | Very difficult | Not applicable |
| 9. **How helpful did you find the information on how to use the oximeter?** *(please circle one response)* | Very helpful | Helpful | Neutral | Not very helpful | Not at all helpful | Not applicable |

| | Yes | No | Not applicable |
|---|---|---|---|
| **10. Did you have anyone (such as a family member or friend) who could help you to use the oximeter if needed?** *(please tick)* | ☐ | ☐ | ☐ |

| | Paper diary | Digital app |
|---|---|---|
| **11. How were you asked to record your symptoms and outcomes** *(please select all that apply)* | ☐ | ☐ |

| | Yes | No |
|---|---|---|
| **12. Were you given the choice between using a digital app or a paper diary?** *(please tick)* | ☐ | ☐ |
| **13. Did you use the same method of recording your symptoms and outcomes throughout?** *(please tick)* | ☐ | ☐ |

| | Yes | No | Not applicable |
|---|---|---|---|
| **14. Were you happy to record using a paper diary?** *(please tick. If you didn't use a paper diary, please choose not applicable)* | ☐ | ☐ | ☐ |

**15. How did you find using the paper diary?** *(please circle one response. If you didn't use a paper diary, please choose not applicable)*

| Very easy | Easy | Neutral | Difficult | Very difficult | Not applicable |
|---|---|---|---|---|---|

| | Yes | No | Not applicable |
|---|---|---|---|
| **16.Did you have access to a smartphone (or other device) to use the digital app?** *(please tick. If you didn't use an app please choose not applicable)* | ☐ | ☐ | ☐ |
| **17.Were you happy to record using a digital app?** *(please tick. If you didn't use an app please choose not applicable)* | ☐ | ☐ | ☐ |

**18.How did you find using the digital app?** *(please circle one response. If you didn't use an app please choose not applicable)*

| | | | | | |
|---|---|---|---|---|---|
| Very easy | Easy | Neutral | Difficult | Very difficult | Not applicable |

| | Yes | No | Not applicable |
|---|---|---|---|
| **19.Did you have anyone to support you with taking and recording readings, if needed?** *(please tick)* | ☐ | ☐ | ☐ |

| | Yes | No | Not sure | Not applicable |
|---|---|---|---|---|
| **20. Did you receive any medications as part of the COVID care at home service?** *(this question may only be applicable for those who have been discharged from hospital)* | ☐ | ☐ | ☐ | ☐ |
| **21. Were you given oxygen as part of the COVID care at home service?** *(this question may only be applicable for those who have been discharged from hospital)* | ☐ | ☐ | ☐ | ☐ |

**22. On average, how often did you (or a family member/friend) speak to a member of the COVID care at home team while receiving the service?** *(please circle one response)*

| | | | | | |
|---|---|---|---|---|---|
| Several times a day | Once a day | Several times a week | Once a week | Less than once a week | Not at all |

**23. How would you rate your contact with the COVID care at home team?** *(please circle one response)*

| | | | | | |
|---|---|---|---|---|---|
| Excellent | Good | Neutral | Poor | Very poor | Not applicable |

**24. What activities were you asked to do as part of your COVID care at home?**
*(Please select all that apply)*

- ☐ Take readings using an oximeter
- ☐ Fill in a diary
- ☐ Provide readings over the phone
- ☐ Provide readings via text
- ☐ Provide readings via email
- ☐ Record readings in a digital app
- ☐ Check over your readings for any issues
- ☐ Seek further help due to a reading being lower than recommended threshold
- ☐ Other (please specify): ______

| Question | | | | | | |
|---|---|---|---|---|---|---|
| **25. Please rate your experience of receiving COVID care at home** *(please circle one option)* | Excellent | Good | Neutral | Poor | Very poor | Not applicable |
| **26. How reassured were you by receiving COVID care at home?** *(please circle one option)* | Very reassured | Reassured | Neutral | Not very reassured | Not at all reassured | Not applicable |

**27. Thinking about your COVID symptoms, how helpful did you find receiving COVID care at home?** *(please circle one option)*

| Very helpful | Helpful | Neutral | Not very helpful | Not at all helpful | Not applicable |
|---|---|---|---|---|---|

**28. Would you recommend COVID care at home to your friends and family** (if in a similar situation) *(please tick)*

☐ **Yes** ☐ **No** ☐ **Not sure**

**How have you found doing the following in practice?** *(please circle one option for each question)*

| | | | | | | |
|---|---|---|---|---|---|---|
| **29.** Using the oximeter | Very easy | Easy | Neutral | Difficult | Very difficult | Not applicable |
| **30.** Recording your readings using the app or diary | Very easy | Easy | Neutral | Difficult | Very difficult | Not applicable |
| **31.** Providing readings to the service | Very easy | Easy | Neutral | Difficult | Very difficult | Not applicable |
| **32.** Seeking further help if have concerns about health (escalating care if necessary) | Very easy | Easy | Neutral | Difficult | Very difficult | Not applicable |
| **33.** Contacting a healthcare professional (if needed) | Very easy | Easy | Neutral | Difficult | Very difficult | Not applicable |
| **34.** Returning the oximeter following discharge | Very easy | Easy | Neutral | Difficult | Very difficult | Not applicable |

| | **Yes** | **No** | **Not sure** |
|---|---|---|---|
| **35. Did you understand what would happen after being discharged from COVID care at home?** *(please tick)* | ☐ | ☐ | ☐ |
| **36. Were you asked to return the oximeter once discharged from COVID care at home?** *(please tick)* | ☐ | ☐ | ☐ |

**37.Did you experience any problems with any of the following?** *(Please select all that apply)*

- ☐ Using the oximeter
- ☐ Recording your readings in the app or diary
- ☐ Providing readings to the service
- ☐ Seeking further help if you had concerns about your health
- ☐ Contacting healthcare professionals when needed
- ☐ Returning the oximeter following discharge
- ☐ Other (please specify):
- ☐ I did not experience any problems

| | Yes | No | Not sure | Not applicable |
|---|---|---|---|---|
| **38.Did you discuss these problems with a member of the COVID care at home team?** *(please tick)* | ☐ | ☐ | ☐ | ☐ |

| | Yes | No | Not sure | Not applicable |
|---|---|---|---|---|
| **39.Was this problem (or were these problems) resolved?** *(please tick)* | ☐ | ☐ | ☐ | ☐ |

**40. What helped or encouraged you to use COVID care at home?** *(Please select all that apply)*

- ☐ Knowing why the service is important (e.g. for patients and/or NHS)
- ☐ Knowing what to do (e.g. how to use the oximeter, how to record readings, etc)
- ☐ Own health (e.g. other health conditions)
- ☐ Having sufficient time
- ☐ Support from family or friends to use the equipment
- ☐ Support from healthcare professionals
- ☐ Wanting to do it
- ☐ Remembering to do it
- ☐ Positive views of the service (e.g. liking it/trust)
- ☐ Other (please specify)
- ☐ None of the above

**41. Was there anything that made it difficult for you to use COVID care at home?** *(Please select all that apply)*

- ☐ Not knowing why the service is important
- ☐ Not knowing what to do (e.g. how to use the oximeter, how to record readings, etc)
- ☐ Own health (e.g. other health conditions)
- ☐ Not having time
- ☐ Not having the right equipment
- ☐ Lack of support from family or friends
- ☐ Lack of support from healthcare professionals
- ☐ Forgetting to do it
- ☐ Not wanting to do it
- ☐ Negative views towards the service (e.g. disliking it)
- ☐ Concerns about being monitored or data use
- ☐ Other (please specify):
- ☐ None of the above

**42. What could have been changed to make it easier for you to engage with COVID care at home?** *(Please describe in the box below, or leave blank if not applicable)*

**43. Which of the following scenarios best describes your experience while receiving COVID care at home?** *(Please tick all that apply)*

- ☐ Stayed at home the whole time
- ☐ Asked to go to the emergency department
- ☐ Admitted to hospital

**44. Is there anything else you'd like to tell us about your experience of receiving COVID care at home?** *(Please describe in the box below, or leave blank if not applicable)*

#### Section D. About you

Finally, we would like to ask you some questions about yourself. If you are completing this survey as a carer, we would also like to ask you some questions about your family member or friend (Section E).

**45. What is your gender?** *(please tick one option)*

☐ Male ☐ Other (please specify)

☐ Female ☐ Prefer not to say

**46. How old are you?** *(please circle one option)*

| | | | | | | | | | | | | | | |
|---|---|---|---|---|---|---|---|---|---|---|---|---|---|---|
| 18-24 years | 25-29 years | 30-34 years | 35-39 years | 40-44 years | 45-49 years | 50-54 years | 55-59 years | 60-64 years | 65-69 years | 70-74 years | 75-79 years | 80-84 years | 85+ years | Prefer not to say |

**47. How many people do you live/co-habit with?** *(please circle one option)*

| | | | | | | | | | | | |
|---|---|---|---|---|---|---|---|---|---|---|---|
| 0 | 1 | 2 | 3 | 4 | 5 | 6 | 7 | 8 | 9 | 10 or more | Prefer not to say |

**48. Which of these best describes your living arrangement?**

☐ I own my home outright

☐ I own my home with a mortgage

☐ I rent from local authority/housing association

☐ I rent privately

☐ Other (e.g. living with friends or family), please specify ____________________

☐ Prefer not to say

**49. What is your ethnic group?** (*Choose one option that best describes your ethnic group or background)*

| **White** | **Asian/Asian British** | **Other ethnic group** |
|---|---|---|
| ☐ English/Welsh/Scottish/ Northern Irish/British<br>☐ Irish<br>☐ Gypsy or Irish Traveller<br>☐ Any other white background, please describe: | ☐ Indian<br>☐ Pakistani<br>☐ Bangladeshi<br>☐ Chinese<br>☐ Any other Asian background, please describe: | ☐ Arab<br>☐ Any other ethnic group, please describe: |
| **Mixed/Multiple ethnic groups**<br>☐ White and Black Caribbean<br>☐ White and Black African<br>☐ White and Asian<br>☐ Any other Mixed/Multiple ethnic background, please describe: | **Black/African/Caribbean/Black British**<br>☐ African<br>☐ Caribbean<br>☐ Any other Black/African/Caribbean background, please describe: | ☐ **Prefer not to say** |

**50. At what age did you complete your continuous full time education?** *(e.g. school, college, further education) Please write your age in the box below:*

____________ years

☐ Prefer not to say

**51. Which of these best describes your highest educational qualification?** *(Please tick one option)*

☐ No formal qualification

☐ Degree level or higher

☐ GCSE/CSE/O level or equivalent

☐ Other, please specify

_________________________

☐ A level/AS level or equivalent ☐ Prefer not to say

**52.Which of these best describes your current work situation?** *(Please select all that apply)*

☐ Working full time
☐ Working part time
☐ Self-employed
☐ Student in higher education
☐ Unemployed
☐ Homemaker
☐ Retired
☐ Furloughed under covid-19
☐ Full time carer (of dependent child or adult)
☐ Not in work due to poor health or disability
☐ Other, please specify ____________________
☐ Prefer not to say

**53.This question is about your sexual orientation. Do you identify as:**

☐ Straight/heterosexual
☐ Gay or lesbian
☐ Bisexual
☐ Other (please specify) [ ]
☐ Prefer not to say

**54.Is English your first language?**

☐ Yes
☐ No, please specify ____________________
☐ Prefer not to say

**55. Prior to your current illness, were your day-to-day activities limited because of a health problem or disability which has lasted, or is expected to last, at least 12 months?**

- ☐ Yes limited a lot
- ☐ Yes limited a little
- ☐ No not limited at all
- ☐ Prefer not to say
- ☐ Not sure
- ☐ Not applicable

**56. What is your postcode?** (*Please write in the box below)*

☐ Prefer not to say

**If you are a family member/friend, please continue to complete Section E (page 16). If you are a patient, please read the summary information below.**

#### Summary

Thank you so much for completing our survey about your experiences of receiving COVID care at home. Your responses will help us to evaluate the COVID care at home service.

Please post your survey responses back to us (the research team) using the pre-paid envelope provided.

Please contact Dr Holly Walton ( |Tel: 020 3108 3068) if you have any questions about this survey.

#### Section E. About your family member/friend [carers only]

If you are completing this survey as a carer, we would also like to ask you some questions about your family member or friend who received COVID care at home. Please complete this section.

If you are a patient, please skip this section and go to the summary section.

**57.What is your relationship with the patient? (***Please tick one option)*

- ☐ Spouse or partner
- ☐ Son or daughter
- ☐ Son or daughter in law
- ☐ Parent
- ☐ Sibling
- ☐ Friend
- ☐ Other, please specify ________________________

**58.How have you supported your family member or friend while they have had COVID?** (*Please provide details in the box below)*

**59.What is the gender of your family member/friend who received COVID care at home?**

- ☐ Male
- ☐ Female
- ☐ Other (please specify)
- ☐ Prefer not to say

**60.How old is your family member/friend?**

| | | | | | | | | | | | | | | |
|---|---|---|---|---|---|---|---|---|---|---|---|---|---|---|
| 18-24 years | 25-29 years | 30-34 years | 35-39 years | 40-44 years | 45-49 years | 50-54 years | 55-59 years | 60-64 years | 65-69 years | 70-74 years | 75-79 years | 80-84 years | 85+ years | Prefer not to say |

**61.What is the ethnic group of your family member/friend?** (*Choose one option that best describes your ethnic group or background)*

| **White** | **Asian/Asian British** | **Other ethnic group** |
|---|---|---|
| ☐ English/Welsh/Scottish/ Northern Irish/British<br>☐ Irish<br>☐ Gypsy or Irish Traveller<br>☐ Any other white background, please describe: | ☐ Indian<br>☐ Pakistani<br>☐ Bangladeshi<br>☐ Chinese<br>☐ Any other Asian background, please describe: | ☐ Arab<br>☐ Any other ethnic group, please describe: |
| **Mixed/Multiple ethnic groups**<br>☐ White and Black Caribbean<br>☐ White and Black African<br>☐ White and Asian<br>☐ Any other Mixed/Multiple ethnic background, please describe: | **Black/African/Caribbean/Black British**<br>☐ African<br>☐ Caribbean<br>☐ Any other Black/African/Caribbean background, please describe: | ☐ **Prefer not to say** |

**62.At what age did your family member/friend complete their continuous full time education?** *(e.g. school, college, further education) Please write your age in the box below:*

____________ years

☐ Prefer not to say

☐ Not sure

**63.Which of these best describes your family member/friend's highest educational qualification?** *(Please tick one option)*

☐ No formal qualification

☐ GCSE/CSE/O level or equivalent

☐ A level/AS level or equivalent

☐ Degree level or higher

☐ Other, please specify ____________________________

☐ Prefer not to say

**64. Which of these best describes your family member/friend's current work situation?** *(Please select all that apply)*

- ☐ Working full time
- ☐ Working part time
- ☐ Self-employed
- ☐ Student in higher education
- ☐ Unemployed
- ☐ Homemaker
- ☐ Retired
- ☐ Furloughed under covid-19
- ☐ Full time carer (of dependent child or adult)
- ☐ Not in work due to poor health or disability
- ☐ Other, please specify ___________________
- ☐ Prefer not to say

**65. This question is about your family member/friend's sexual orientation. Do they identify as:**

- ☐ Straight/heterosexual
- ☐ Gay or lesbian
- ☐ Bisexual
- ☐ Other (please specify) [ ]
- ☐ Prefer not to say

**66. Is English your family member/friend's first language?**

- ☐ Yes
- ☐ No, please specify ___________________
- ☐ Prefer not to say

**67. Prior to your family member/friend's current illness, were their day-to-day activities limited because of a health problem or disability which has lasted, or is expected to last, at least 12 months?**

☐ Yes limited a lot
☐ Yes limited a little
☐ No not limited at all
☐ Prefer not to say
☐ Not sure

**68. What is your family member/friend's postcode?** (*Please write in the box below)*

☐ Prefer not to say

☐ Not sure

#### Summary

Thank you so much for completing our survey about your family member/friend's experiences of receiving COVID care at home. Your responses will help us to evaluate the COVID care at home service.

Please post your survey responses back to us (the research team) using the pre-paid envelope provided.

Please contact Dr Holly Walton ( |Tel: 020 3108 3068) if you have any questions about this survey.
