## Supplementary files 5-8 for "Patients’ and carers’ experiences of, and engagement with remote home monitoring services for COVID-19 patients: a rapid mixed-methods study"

**Appendix 5. Demographic characteristics for patient and carer survey respondents**

|  |  | **n (%)** |
| --- | --- | --- |
| Survey respondent | Patient | 936 (87.6) |
|  | Carer | 48 (4.5) |
|  | Unknown | 85 (8) |
|  | Total | 1069 (100) |

| **Demographic characteristic** | | **Patient**  **n (%)** | **Carer**  **n (%)** |
| --- | --- | --- | --- |
| Gender  (patient n=920; carer n=45) | Female | 531(58) | 27 (60) |
|  | Male | 385 (42) | 18 (40) |
|  | Other/prefer not to say | 4 (0.4) | 0 |
| Age  (patient n=923; carer n=46) | Under 50 years | 195 (21.1) | 13 (28.3) |
|  | 50-64 years | 428 (46.4) | 24 (52.2) |
|  | 65-79 years | 256 (27.8) | 4 (8.7) |
|  | >=80 years | 43 (4.7) | 5 (10.9) |
|  | Prefer not to say | 1 (0.1) | 0 |
| Living circumstances (patient n=863) | Living alone | 132 (15.3) |  |
|  | Household of 2 | 339 (39.3) |  |
|  | Household of 3 | 152 (17.6) |  |
|  | Household of 4/5/ | 201 (23.3) |  |
|  | Household of 6+ | 36 (4.2) |  |
|  | Prefer not to say | 3 (0.3) |  |
| Ethnicity  (patient n=918; carer n=47) | White British/English/Welsh/Scottish/ Irish or any other white background  Black/African/Caribbean/Black British or any other Black background  Asian/Asian British or any other Asian background  Mixed or multiple ethnic background  Any other ethnic group  Prefer not to say | 836 (91.1)  16 (1.7)  48 (5.2)  12 (1.3)  2 (0.2)  4 (0.4) | 38 (80.9)  0  9 (19.1)  0  0  0 |
| Highest educational qualification  (patient n=914; carer n=46) | No formal qualification  GCSE/CSE/O level or equivalent  A level/AS level or equivalent  Degree level or higher  Other  Prefer not to say/not sure | 146 (16)  273 (29.9)  106 (11.6)  212 (23.2)  80 (8.8)  97 (10.6) | 10 (21.7)  16 (34.8)  8 (17.4)  7 (15.2)  1 (2.2)  4 (8.7) |
| Age completed full time education  (patient n=791; carer n=28) | 15 years or under  16 years  17-18 years  19-21 years  >21 years  Prefer not to say | 146 (18.5)  267 (33.8)  163 (20.6)  104 (13.1)  99 (12.5)  12 (1.5) | 4 (14.3)  7 (25.1)  12 (42.9)  2 (7.1)  3 (10.7)  0 (0.0) |
| Work situation*  (patient n=969; carer n=48) | Working full time  Working part time  Self-employed  Student in higher education  Unemployed  Homemaker  Retired  Furloughed  Full time carer  Not in work due to poor health or disability  Other/prefer not to say | 355 (37.6)  128 (13.5)  41 (4.3)  2 (0.2)  18 (1.9)  21 (2.2)  274 (29)  15 (1.6)  19 (2)  65 (6.9)  31 (3.3) | 16 (33.3)  6 (12.5)  1 (2.1)  0  3 (6.3)  3 (6.3)  9 (18.8)  0  1 (2.1)  5 (10.4)  1 (2.1) |
| Sexual orientation  (patient n=919; carer n=44) | Straight/heterosexual  Gay or lesbian  Bisexual  Other/Prefer not to say | 858 (93.4)  13 (1.4)  5 (0.5)  43 (4.7) | 41 (93.2)  0  0  3 (6.8) |
| English as first language (patient n=925; carer n=43) | Yes  No  Prefer not to say | 852 (92.1)  66 (7.1)  7 (0.8) | 35 (81.4)  8 (18.6)  0 |
| Day to day activities limited by a health problem or disability (patient n=920; carer n=46) | Limited a lot or a little  Not limited at all  Prefer not to say  Not sure/not applicable | 351 (38.1)  482 (52.4)  4 (0.4)  83 (9) | 20 (43.4)  17 (37)  1 (2.2)  8 (17.4) |
| Deprivation score (patient n=767; carer n=37) | Q1 or Q2 (Most deprived)  Q3 or Q4  Q5 or Q6  Q7 or Q8  Q9 or Q10 (Least deprived) | 182 (23.7)  137 (17.9)  149 (19.4)  161 (21)  138 (18) | 13 (35.1)  5 (13.5)  7 (18.9)  9 (24.3)  3 (8.1) |
| Relationship to patient  (carer n=42) | Spouse or partner  Son or daughter  Other |  | 24 (57.1)  11 (26.2)  7 (16.7) |

*Respondents able to select more than one option

**Appendix 6.** Demographic characteristics for patient and carer interview respondents

| **Demographic characteristic** | | **Patient**  **N (%)*** | **Carer**  **n (%)** |
| --- | --- | --- | --- |
| Patient or carer who took part in the interview |  | 59 (95%) | 3 (5%) |
| Gender | Female | 31 (50%) | 3 (100%) |
|  | Male | 31 (50%) | 0 |
| Age | Under 50 years | 8 (13%) | 2 (67%) |
|  | 50-64 years | 31 (50%) |  |
|  | 65-79 years | 21 34% | 1 (33%) |
|  | >=80 years | 2 (3%) |  |
| Living circumstances | Live alone | 5 (8%) |  |
|  | Household of 2 | 36 (58%) | 2 (67%) |
|  | Household of 3 | 11 (18%) |  |
|  | Household of 4-5 | 9 (15%) |  |
|  | Household of 6+ | 1 (2%) | 1 (33%) |
| Home ownership/renting | Own home outright | 26 (42%) | 1 (33%) |
|  | Own home with mortgage | 16 (26%) | 1 (33%) |
|  | Own home (not specified) | 2 (3%) |  |
|  | Rent from local authority/house association | 9 (15%) |  |
|  | Rents privately | 6 (10%) |  |
|  | Other | 2 (3%) | 1 (33%) |
| Ethnicity | White British/English/Welsh/Scottish  White Irish  Any other white background | 50 (81%) |  |
|  | Black/African/Caribbean/Black British | 3 (5%) |  |
|  | Asian/Asian British | 7 (11%) | 2 (67%) |
|  | Missing | 1 (2%) | 1 (33%) |
|  | Not enough information | 1 (2%) |  |
| Age completed full time education of | 15 years or under | 13 (21%) |  |
|  | 16 years | 21 (34%) |  |
|  | 17-18 years | 16 (26%) |  |
|  | 19-21 years | 5 (8%) |  |
|  | >21 years | 4 (6%) | 1 (33%) |
|  | Not known | 2 (3%) | 2 (67%) |
| Highest educational qualification | No formal qualification | 14 (23%) |  |
|  | GCSE/CSE/O level or equivalent | 21 (34%) |  |
|  | A level/AS level or equivalent | 5 (8%) |  |
|  | Degree level or higher | 14 (23%) | 2 (67%) |
|  | Other | 7 (11%) |  |
|  | Not sure | 1 (2%) | 1 (33%) |
| Work situation | Working full time | 23 (37%) | 2 (67%) |
|  | Working part time | 1 (2%) |  |
|  | Self-employed | 2 (3%) |  |
|  | Not working | 1 (2%) |  |
|  | Homemaker | 2 (3%) |  |
|  | Retired | 22 (36%) |  |
|  | Furloughed | 1 (2%) |  |
|  | Not in work due to poor health or disability | 7 (11%) |  |
|  | Not sure | 2 (3%) | 1 (33%) |
| Sexual orientation | Straight/heterosexual | 62 (100%) | 3 (100%) |
| English as first language | Yes | 54 (87%) | 3 (100%) |
|  | No | 7 (11%) |  |
|  | Not specified | 1 (2%) |  |
| Day to day activities limited by a health problem or disability | Yes limited a lot | 6 (10%) |  |
|  | Yes limited a little | 6 (10%) |  |
|  | Limited (but not specified how much) | 4 (6%) |  |
|  | No not limited at all | 46 (89%) | 2 (67%) |
|  | Not known |  | 1 (33%) |
| Relationship to patient | Spouse or partner |  | 1 (33%) |
|  | Son or daughter |  | 2 (67%) |
| Note. *59 patients took part in the interviews, but we have demographic characteristics for 62 patients as carers reported patient demographics too | | | |

**Appendix 7.** Example findings for factors influencing engagement.

| **Theme** | **Sub-theme** | **Key findings (interviews and survey)** | **Example quotes (illustrative of key findings for each sub-theme)** |
| --- | --- | --- | --- |
| Patient factors | Knowledge | **Facilitators**   - 55% (n=583/1069) of respondents felt that knowing what to do helped them to engage with the service *(survey findings)* - Many participants felt that they had the appropriate knowledge to use the oximeter, monitor and record/communicate readings and escalate care. *(interview findings)*   **Barriers**   - 1% (n=7/1069) reported that not knowing what to do was a barrier to engagement *(survey findings)* - Some participants mentioned in ‘other’ that not knowing what to do in regards to escalating care was a barrier *(survey findings)* - Some survey participants mentioned problems in relation to not knowing what to do following discharge *(survey findings)* - Interview findings indicated that one of the main barriers was a lack of knowledge for all pathway aspects. This included: a lack of knowledge in relation to understanding and interpreting information and equipment, language barriers, a lack of knowledge on how to fill out the diary or complete the recordings, knowledge barriers around escalating care and knowing when to call for help *(interview findings)* | - *“Well I mean. It’s quite straightforward isn’t it. You just put it on your finger and let it settle down and read the figures off. Pulse-pulse and oxygen levels. So no, I didn’t find it complex at all”* (Site C, interviewee 2) - *“I’ll be truthful, no, not to tell you now, however, I’m sure in the instructions, it would have been there but no I couldn’t tell you what they meant, I just recorded it and I think when you recorded it in the app, I think that’s where you would trust the doctors or the nurses to phone and say, “That stat doesn’t feel right”.* (Site I, interviewee 6) - *“And I think my dad got confused a couple of times with the readings, because she would still be asleep when he’d do the observations. And obviously you’re not supposed to do when they’re asleep”* (Site F, interviewee 6) |
|  | Memory | **Facilitators**   - Some participants spoke about how phone calls helped them to remember to do the readings or that they wrote their readings on post it notes to facilitate memory *(interview findings)*   **Barriers**   - 4% (n=37/1069) reported that forgetting to do it was a barrier to engaging with the service *(survey findings)* - Memory was also reported as an issue for monitoring and recording and communicating readings *(interview findings)* | - *“I think it was better to actually take the readings while they were talking to me on the phone, because the majority of people possibly around my age and probably a little bit older, their memory probably wouldn’t be as good, and they could get their figures distorted and so forth if they had to write them down. So the way it was approached over the phone was more reliable”* (Site N, interviewee 1) - *“And eventually I seemed to reach the point that I hadn’t done it for so long that I got a phone call, but that was just by me being really forgetful rather than wanting to waste people’s time”* (Site N, interviewee 3) |
|  | Physical health | **Facilitators**   - 54% (n=581/1069) of respondents felt that their own health helped them to engage with the service *(survey findings)*   **Barriers**   - 4% (n=39/1069) of respondents reported that their own health was a barrier to engagement, and some participants mentioned in ‘other’ that being too unwell to take readings or use app was a barrier *(survey findings)* - Barriers relating to physical health included: feeling too poorly/not in the right frame of mind / sleeping a lot / having a health condition that made it difficult to monitor / needing to ask people to repeat themselves / difficulties getting to telephone / difficulties reading oximeter due to eyesight *(interview findings)* | - *“Sometimes the reading took quite a long time to come through and that was because of my circulation, my circulation was quite poor so it was taking a while to calibrate”* (Site B, interviewee 2) - *“I’ve got a severe visual impairment, so I couldn’t see the numbers and everything, so that had to do all the for me” (Site E, interviewee 4)* - *“Really they just, in the main I was quite poorly, in fact I would say I was really poorly, it’s the only time I’ve thought I was going to die in my life […] so really in the main my husband dealt with them, I couldn’t really be remotely bothered with them if I’m honest and I can’t remember what they told me, I don’t think they told me a lot” (Site I, interviewee 3)* |
|  | Attitudes towards the service and behaviours | **Facilitators**   - 60% (n=637/1069) of respondents felt that knowing why the service is important helped them to engage with the service *(survey findings)* - 46% (n=491/1069) of respondents felt that wanting to do it helped them to engage with the service *(survey findings)* - 27% (n=288/1069) felt that positive views of the service helped them to engage with the service, some participants mentioned in ‘other’ that the support/reassurance helped them to engage *(survey findings)* - Determination, knowing it was important, being happy to follow advice, liking contact via phone, motivation, seeing their own progress and confidence escalating care facilitated engagement *(interview findings)*   **Barriers**   - 1% (n=8/1069) felt that concerns about being monitored or data use was a barrier to engagement *(survey findings)* - 1% (n=6/1069) felt that not wanting to do it was a barrier to engagement *(survey findings)* - A key barrier was a lack of interest in monitoring/recording, or views that monitoring did not help or that it made the participant anxious if the reading was low *(interview findings)* - Barriers to seeking further support/escalating care included worries and concerns around going to hospital (due to COVID-19, lack of support in hospital, difficulties communicating, covid risk) *(interview findings)* | - *“But there are sometimes, I must admit, sometimes it makes you feel a little bit anxious [..] but then you can leave it – because you want to get things all right – but towards the end of the day it is so worth it – it is 100% worth it to have the oximeters reading every day to know; to understand where you are […] but I still guarantee that 100% it is a very good idea”* (Site A, interviewee 1) - *“Just really, they encouraged me to ring an Ambulance if I needed it. And I wasn’t ringing them, because I felt like I was wasting their time, or whatever. I didn’t want to, because I was worried they might want to take me in”* (Site B, interviewee 4) - *“And it came to a point where her oxygen levels had dropped significantly that morning. I rang the nurse. She was like, you need to get an ambulance to your mum. And we were like, look we really don’t want to send her to hospital”* (Site F, interviewee 6) |
|  | Time to do behaviours | **Facilitators**   - 20% (n=217/1069) of respondents felt that having sufficient time helped them to engage with the service *(survey findings)* - Building the behaviours into their routine and keeping equipment nearby made it easier *(interview findings)*   **Barriers**   - lack of time (those working from home) made it difficult *(interview findings)* | - *“To be quite honest after I got used to exactly what it was asking me, I was then ready for exactly each thing. I was getting through the phone call within less than a couple of minutes”* (Site B, interviewee 1) - *“No, no it was all really clear and after the first couple of days it became like a routine” (Site J, interviewee 2)* - *“Well it probably says more about me than anything else, but there was times when I was catching up, even though I had nothing else much to do, I was trying to busy myself with activities, even if it was just watching a silly video […]. So even though I was just sat there not really doing anything, I would still think, Oh my gosh, it’s 10 o’clock, I haven’t done my 9 o’clock […] I’m really bad with time keeping. So you’d think, just set an alarm and do it. Which is exactly what I did […] and that solved that problem”* (Site N, interviewee 3) |
| Wider support and resources | Support from staff/service | **Facilitators**   - 46% (n=488/1069) of respondents felt that support from healthcare professionals helped them to engage with the service, some participants mentioned in ‘other’ that a clinician referring them helped them to engage *(survey findings)* - Support from staff/service was a facilitator: it helped to understand information, helped with monitoring e.g. dropping oximeter off, being responsive to questions about monitoring, with recording & communicating e.g. ringing patients if they forgot, being patient/giving feedback, face to face appointments, continuity and explanations. And support in escalating care – having a number to call/positive views/practical advice/reassurance *(interview findings)*   **Barriers**   - 2% (n=24/1069) reported that lack of support from healthcare professionals was a barrier to engagement, some participants mentioned in ‘other’ that being unable to contact staff was a barrier (survey findings) - A few participants did report lack of support from staff re. recording and communicating readings and also that some participants couldn’t get through to the team when needed *(interview findings)* | - *“The nurse was very good, can’t praise her really high enough. She was a friendly voice to speak to. Fair enough, you know, I’ve got a bit of a support system, but for somebody who hasn’t got that much of a support system around them, I think that friendly voice would go a long way just to, you know, easing their minds” (Site D, interviewee 5)* - *“And then near the end when I was getting a bit complacent, I was sort of almost well, a couple of times I didn’t put them in and they would phone and say, “Are you okay? You’ve not submitted you reading.” So that was just really supportive, and I said it certainly reassured me”* (Site M, interviewee 1) - *“So, the lady phoned me in the morning to say that the machine would be dropped off and then she said that they would text me every day and i said “Are you not coming out to see me?” and she said, “No, we will text you and we are on the end of the phone.” […] I thought if they are coming out to see me, they will know by looking at me or if I’m okay but nobody ever came out [..] because you feel like you are just on your own, just left on your own really”* (Site D, interviewee 3) |
|  | Support from family members/friends | **Facilitators**   - 25% (n=266/1069) of respondents felt that support from family members/friends helped them to engage with the service (survey findings) - Support from family/members and friends helped with all aspects of the process including understanding information, collecting oximeter and other equipment, doing the monitoring, recording and communicating readings, escalating care *(interview findings)* - Prior family experience was also found to be helpful *(interview findings)*   **Barriers**   - 1% (n=9/1069) reported that lack of support from family and friends was a barrier to engagement *(survey findings)* | - *“So my dad was initially involved in I think it was nine days, so the first nine days he took full care of mum to be honest clinically I was involved in a lot of the calls because I think my dad’s getting quite stressed. […] so yes, he did the physical side of it. He would do the observations. And then he’d call me first thing in the morning, or he’d drop a text to say these are the observations. I’d call and have a quick chat, knowing the nurse was going to call us. So I guess it was a bit of a joint effort between us”* (Site F, interviewee 6) - “My husband had to fill it in, he had to take my temperature and things like that, and they were phoning up every day for the results. And if my husband had any questions he wanted to ask, if he was worried or anything, they told him what to do and if there was any problems” (Site K, interviewee 2) - *“In the beginning I found it difficult because I was poorly but then I had a member of my family to help me and then it was just east to do and I’m not good at things like that. I found it easier to do after a while. It’s just when you’re poorly you find it a bit difficult but no, it was easy. […] they would help me to do my test. They would sit with me and help me write it down and then send it through to the text. That was when I first became, you know, poorly with it and I wasn’t sure what to do and they found it easy to help me” (Site C, interviewee 6).* |
|  | Accessibility and availability of materials | **Barriers**   - Many participants spoke about how they did not receive enough/received too much information and that some information was contradictory and confusing and not accessible *(interview findings)* - Some participants were not given resources such as paper diaries *(interview findings)* | - *“I had a leaflet and everything, but again the only thing with that, because every time I did get a call from the doctor. Quite often it was a different doctor […] I was getting different advice. […] I think the leaflet says to use either your middle finger or your pointy finger. A couple of times I got dodgy readings or a little bit off. So, some of the doctors were saying use your little finger, obviously keep it steady.”* (Site P-A, interviewee 1) |
|  | Equipment | **Facilitators**   - Some participants spoke about buying their own equipment or already having the equipment and that this helped *(interview findings)*   **Barriers**   - 1% (n=8/1069) reported that not having the right equipment was a barrier to engagement, some participants mentioned in ‘other’ that the oximeter not working was a barrier *(survey findings)* - Barriers to monitoring included not being able to collect oximeter, equipment not working and not being able to use own equipment *(interview findings)* | - *“I didn’t have a thermometer, no, so I didn’t know what my temperature was. It never asked you for the temperature anyway. I wouldn’t have been able to do it anyway because I didn’t have one”* (Site A, interviewee 5) |
|  | Technology | **Facilitators**   - Some participants mentioned in ‘other’ that reminder texts or alerts from the app helped them to engage *(survey findings)* - Some participants said that the technology being easy to use helped *(interview findings)*   **Barriers**   - Some participants mentioned in ‘other’ that difficulties with the app was a barrier *(survey findings)* - Some survey participants mentioned problems relating to the oximeter not working, and tech problems *(survey findings)* - Some participants spoke about how technology systems had some barriers e.g. clunky / not including free text *(interview findings)* | - *“I mean perhaps with other did offer text but maybe they were away I don’t have a smart phone, well I don’t have a mobile phone”* (Site B, interviewee 5) - *“I didn’t have any problems. [..] But no, we didn’t have any problems using any of the machinery”* (Site L, interviewee 5) |
| Service factors | Monitoring characteristics | **Barriers**   - Some participants mentioned in ‘other’ that the inconsistent timing of calls was a barrier *(survey findings)* - Some survey participants mentioned problems relating to too many calls *(survey findings)* - Interview findings indicated that monitoring characteristics were a barrier in some cases for example, not being able to see the data to look at progress, monitoring not capturing difficulties and the frequency of monitoring and recording being too much/too frequent *(interview findings)* - Some participants also wanted more phone calls or a more consistent phone call time *(interview findings)* | - *“I found, to start with I found the text messages useful but the longer they went on the more irritating. I was, I felt like I was chained to the phone and you know and to my equipment. So three times a day is, I know that’s necessary to start with but I just felt that maybe twice a day after that might have been better”* (Site B, interviewee 3) - *“Mostly afternoons which were ideal because I’d already done a few readings, a couple of readings. But the thing is the odd time ringing at say 10 o’clock in the morning and because I’d only done one like I’d had to do another one there and then first while I was talking. It would have been better ringing up afternoon type thing so you’ve already done a couple of readings”* (Site F, interviewee 5) |
|  | Service characteristics | **Facilitators**   - Some participants mentioned in ‘other’ that being able to be cared for at home helped them to engage *(survey findings)*   **Barriers**   - Some survey participants mentioned problems relating to a delay in enrolment, and limited hours of service operating (survey findings) - Some participants wanted to continue monitoring after the service *(interview findings)* | - *“I think that lapse in time was because they didn’t realise that I had been discharged, so there was a little bit of miscommunication there I think. But again it’s a new thing isn’t it so*” (Site B, interviewee 1) - *“They were quite easy to contact”* (Site C, interviewee 2) - *“So at the time I wasn’t having any shortness of breath, any light headedness or anything but if that changed then obviously, I’d need to phone them but I didn’t have their number” (Site C, interviewee 4)* |
|  | Scope of service | **Barriers**   - Scope of service was a barrier to escalation as some participants didn’t know whether to ring to ask for help. Additionally others felt the service was not holistic (i.e. did not cover all wider symptoms of COVID) *(interview findings)* | - *“But I think what confused them was that there was – and I wasn’t really dropping below 90 at that time and I think they thought it was COPD so they didn’t you know – that’s my only feedback to all of it. There again just because you’ve got Covid and just because your positive, if you don’t tick the boxes like short of breath, cough and high temp […] it will all unfold in a minute […] I think they probably had a very clear directive as to what they were looking for the objective of this support system yeah. And I sort of slipped – I can’t say slipped through the net”* (Site B, interviewee 2) - *“I think somebody should maybe discuss some of the other things. To me, I got the impression that as long as I as breathing and my oxygen levels were reasonable, that is all they were interested in. Where there were other things that I was a bit concerned about which I don’t think were discussed unless I brought it up”* (Site J, interviewee 1) |
|  | Availability of treatment | **Barriers**   - Some survey participants mentioned problems relating to difficulties contacting their GP *(survey findings)* - Inability to receive oxygen in own home if needed *(interview findings)* | - *“At times I could have done with an extra shot of oxygen I can assure you”* (Site F, interviewee 5) - *“I would just think that this is a service that should be there for everybody”* (Site B, interviewee 4) |

*Appendix 8.* Patient/carer recommendations to improve the service

| **Category** | **Recommendation** |
| --- | --- |
| Patient awareness | - Need more publicity about the service - Need to know about the service sooner or earlier referral - Need to improve link to NHS Track and Trace - Need national standardised approach or automatic referral |
| Patient enablement | - Need more information provision about the service at referral, about escalation and about discharge - Need a dedicated contact number for patients to contact service if needed - Some patients require some face-to-face visits - Need reminders to submit readings |
| Workforce | - More continuity of staff and continuity of information - More contact with doctors desired by some patients |
| Individual differences | - More personalised approach - Flexibility of methods and patient choice |
| Equipment | - Provision of thermometers - Support with using technology (oximeter and digital platforms) |
| Logistics of service | - Consider timing of monitoring calls and having a specific time slot - Need to receive calls when promised - Need to ensure practical and efficient arrangement of oximeter delivery and return |
| Signposting | - Following patients up after discharge - A point of contact after discharge to ask questions to - Signposting to places that can support them whilst on the service - Community drives to ease concerns over hospitals |
